# A spatial multi-omic portrait of survival outcome for clear cell renal cell carcinoma

**DOI:** 10.64898/2026.03.02.26347390

**Authors:** Lasse Meyer, Stefanie Engler, Marlene Lutz, Peter Schraml, Dorothea Rutishauser, Anne Bertolini, Matthias Lienhard, Christian Beisel, Franziska Singer, Natalie De Souza, Niko Beerenwinkel, Holger Moch, Bernd Bodenmiller

## Abstract

Clear cell renal cell carcinoma (ccRCC) is the leading cause of kidney cancer-related death, but how the tumor microenvironment shapes patient survival is not completely understood. Here, we describe the characterization of ccRCC tumor ecosystems from 498 patients using imaging mass cytometry with a focus on tumor, myeloid, and T cell landscapes. Data from more than 3 million single cells is analyzed using machine-learning to identify key ecosystem features that outperform basic clinical data for predicting patient survival. We define three survival ecotypes of ccRCC: *Poor* ecotypes, correlate with the worst survival, have high levels of ICAM1 and CD44 expression in tumor cells and are enriched in M2-like macrophages and interactions of exhausted CD8^+^ T cells with macrophages. *Favorable* ecotypes are characterized by high levels of VHL on tumor cells and of HLADR on myeloid cells and contain Th1-like CD4^+^ T cells. *Medium* ecotypes have the highest endothelial cell density and various immune-to-tumor interactions. Multi-omic characterization of these ecotypes using targeted genomic sequencing and metabolic imaging reveals distinct genomic and metabolic features, including *BAP1* mutations in *Poor* and *VHL* monodriver/wild-type status in *Favorable* patients. We show that deep learning allows ecotype prediction directly from standard pathology H&E images. We validate the ecotypes and their associated molecular characteristics with orthogonal omics data across five clinical cohorts and more than 2,500 patients. These analyses highlight an overall survival benefit for *Medium* patients treated with immunotherapy. In summary, our study distills the survival-relevant information encoded in the ccRCC tumor microenvironment into prognostic survival ecotypes, which may inform clinical decision making in the future.

## INTRODUCTION

Renal cell carcinoma (RCC) is one of the ten most common cancers worldwide and has a rising global incidence. RCC mainly presents as clear cell RCC (ccRCC), the leading cause of kidney cancer-related death^1,2^. Treatment success depends on disease stage, and a large proportion of patients are diagnosed with advanced and metastatic disease. Although targeted and immune checkpoint inhibitor therapies have significantly improved patient outcomes, most patients do not have a sustained response to treatment^3–6^. Validated clinical biomarkers for therapeutic response are scarce, and none are available that correlate with response to immunotherapy^7^.

The tumor microenvironment (TME) of ccRCC is a key player in this heterogenous clinical picture. Single-cell studies based on single-cell RNA sequencing (RNA-seq) and mass cytometry data have highlighted phenotypic shifts in the tumor, T, and myeloid cell compartments to be important in disease progression and immunotherapy response, including distinct tumor states and presence of exhausted CD8^+^ T cells and immunosuppressive macrophages^8–14^. However, studies to date have profiled a small number of patients and typically lack spatial resolution. An assessment of how the single-cell and spatial complexity of the TME impacts patient survival in ccRCC is therefore still missing.

Genomic alterations and metabolic deregulation are hallmarks of ccRCC^15,16^. Aberrant activation of signaling mediated by the hypoxia-inducible factor HIF, due to near universal loss of chromosome 3p genes and inactivation of the *VHL* gene, and additional mutations in the chromatin-remodeling genes *PBRM1*, *BAP1*, and *SETD2* direct tumor evolution, drive metabolic complexity, and impact patient and immunotherapy outcomes^17–22^. Molecular subtypes of ccRCC with different genomic, transcriptomic, proteomic, and metabolic traits and distinct survival patterns have been proposed but have not been translated to guide clinical decision making^23–27^. Moreover, these studies were largely based on bulk tumor profiling and thus cannot be correlated with single-cell and spatial features of the TME.

Here, we conducted a detailed analysis of how the survival of ccRCC patients is shaped by tumor, T, and myeloid cell landscapes as well as by genomic, metabolic and morphology features. We comprehensively characterized the ccRCC ecosystems of tumors from 498 patients using imaging mass cytometry (IMC)^28^, targeted genomic sequencing, and metabolic analyses performed with MALDI mass spectrometry imaging (MALDI-MSI). Our machine- and deep-learning based analyses identified key survival-relevant ecosystem features that outperformed standard clinical data for survival prediction and defined ccRCC survival ecotypes with specific genomic and metabolic attributes. We show that these survival ecotypes can be predicted from standard pathology hematoxylin and eosin (H&E) stained images. We validated our findings by analyses of orthogonal omics data from five independent ccRCC patient cohorts including around 2,500 patients; this analysis revealed distinct treatment avenues beneficial for each ecotype.

## RESULTS

### Highly multiplexed imaging of the ccRCC ecosystem

To create a multi-modal spatial map of survival-relevant ecosystem features for ccRCC, we employed single-cell resolution IMC and complementary modalities, spanning proteomic, morphological, genomic, metabolomic, and transcriptomic data **(Figure 1A)**. For IMC, we analyzed tumor microarrays that included expert pathologist-selected pre-treatment tumor areas from 498 ccRCC patients for whom extensive clinical information, including overall survival (OS) with more than 10 years of follow-up, T stage, ISUP grade, and necrosis status, were available **(Figure 1B)**. Patient survival in this cohort followed expected trends **(Figure 1C, D)**. Consecutive tissue sections were stained with three IMC antibody panels tailored to characterize tumor, T, or myeloid cells **(Figure 1E and Table S1)**. These panels were designed to detect distinct phenotypic profiles of each compartment of interest^10^, cellular states, and immunoregulatory proteins as well as general cell-type markers of the TME. 85 unique targets were profiled in 3,211,104 single cells across 2924 images (up to 6 images per patient per panel) **(Figure S1A)**. This generated, to our knowledge, the largest spatially resolved single-cell dataset of ccRCC analyzed to date.

**Figure 1:**
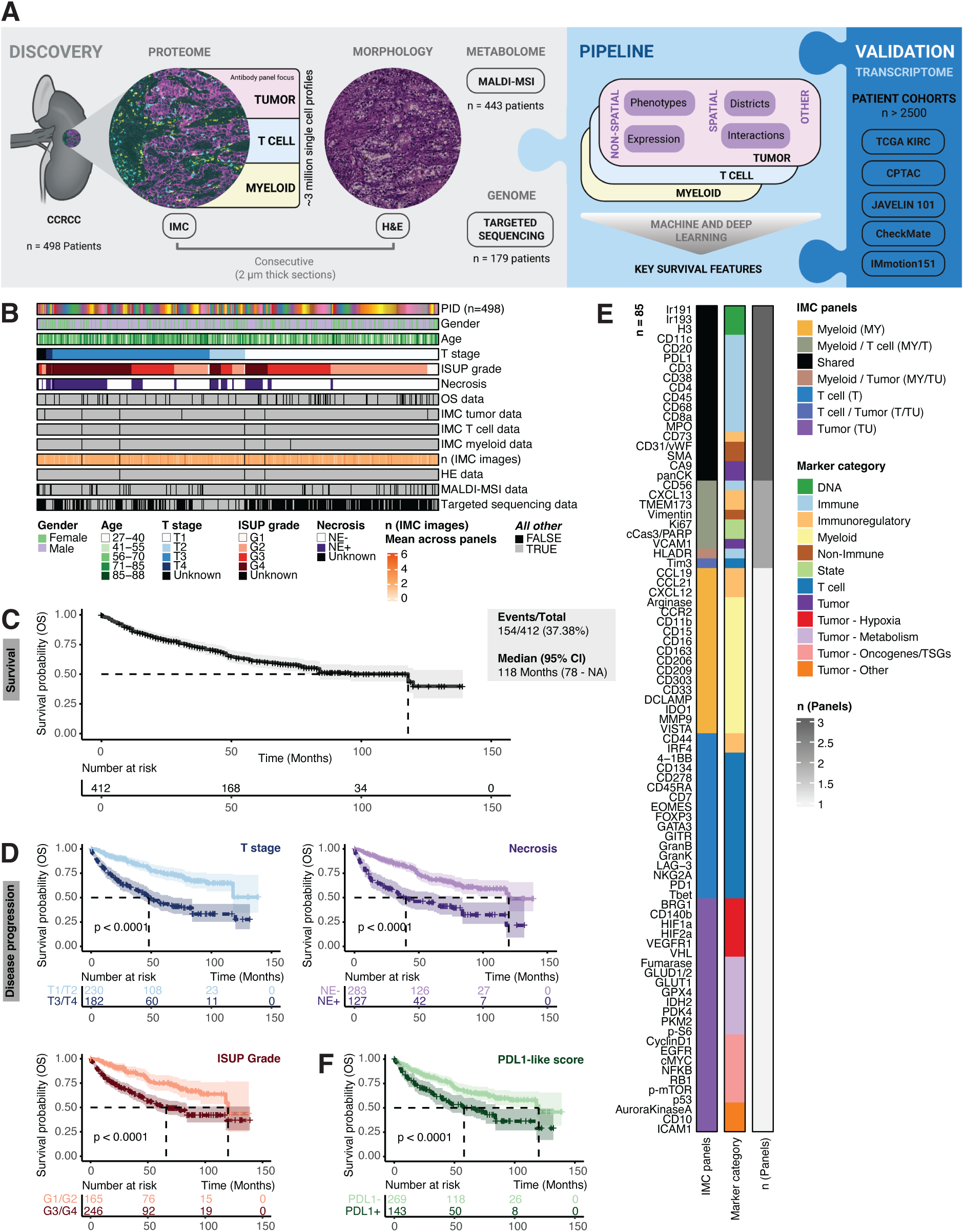
High-dimensional imaging of the ccRCC ecosystem. *See also **Figure S1**.* ***(A)*** Experimental and analytical workflow (created with BioRender.com). ***(B)*** Map of available clinical metadata for the discovery cohort (n=498 patients) ordered by T stage (size and extent of primary tumor), ISUP grade (nucleolar prominence and presence of nuclear anaplasia, giant cells, rhabdoid, and sarcomatoid features), and necrosis (presence or absence). PID, patient ID; OS, overall survival. ***(C)*** Kaplan-Meier curves of OS (n=412 patients). Only patients for whom IMC data were available for all three panels are shown. ***(D, F)*** Kaplan-Meier curves of OS stratified into groups by ***D)*** T stage, ISUP grade, and necrosis and ***F)*** PDL1-IMC score. *P* values were estimated using a log-rank test. ***(E)*** Overview of markers targeted by the antibodies in the three IMC antibody panels used in this study. TSGs, tumor suppressor genes.

For downstream analysis of data from each panel, we split tumor and non-tumor cells and assigned non-tumor cells to broad TME cell types (i.e., B cells, CD4^+^ and CD8^+^ T cells, plasma cells, macrophages, neutrophils, endothelial cells, and stromal cells) using a combination of unsupervised clustering, Gaussian mixture modeling, and deep learning prediction^29^ based on expression of key markers **(Figure S1B, C and Table S2)**. Assigned cell types had expected marker expressions, and their densities were well correlated across panels and patients **(Figure S1D-F)**. Further, we classified each patient based on whether or not ≥1% of cells were PDL1^+^ based on our IMC data. Those subjects with ≥1% PDL1^+^ cells had poorer prognosis, and this PDL1-IMC score served as a proxy for immune-related disease progression **(Figure 1F and Figure S1G)**.

### Ecosystem features are associated with survival and disease progression

As a first step toward a systematic understanding of which TME features are predictive of clinical outcome in ccRCC, we analyzed the tumor, T cell, and myeloid cell compartments in detail with two non-spatial metrics, expression (i.e., cell-type specific marker levels) and phenotypes (i.e., cellular profiles derived from unsupervised clustering of expression patterns), and two spatial metrics, districts (i.e., homotypic spatial aggregations of cell phenotypes in close proximity **(Figure S2A)**) and interactions (i.e. heterotypic cell-to-cell interaction patterns of cell phenotypes). We then used a combination of multivariable Cox proportional hazards modelling and Kaplan-Meier survival estimates to identify those features associated with OS.

For tumor cells, expression of CD44, ICAM1, and PDL1 was associated with worse prognosis, whereas expression of VHL, CyclinD1, NFKB, and p-mTOR, among others, was positively prognostic **(Figure 2A)**. We identified five prognostic tumor phenotypes using unsupervised clustering of cell type-relevant markers **(Figure 2A and Figure S2B)**. For example, the ICAM1^hi^ CD73 phenotype was related to bad prognosis, and the HLADR^hi^ phenotype was related to good prognosis. Spatially, and in keeping with our non-spatial results, we observed that districts dominated by ICAM1-expressing tumor cells were related to poor prognosis, whereas districts with high p-mTOR tumor expression were related to favorable prognosis **(Figure 2A and Figure S2C)**. We also found that interactions of tumor cell phenotypes with endothelial cells were positively prognostic, but interactions with macrophages associated with worse prognosis.

**Figure 2:**
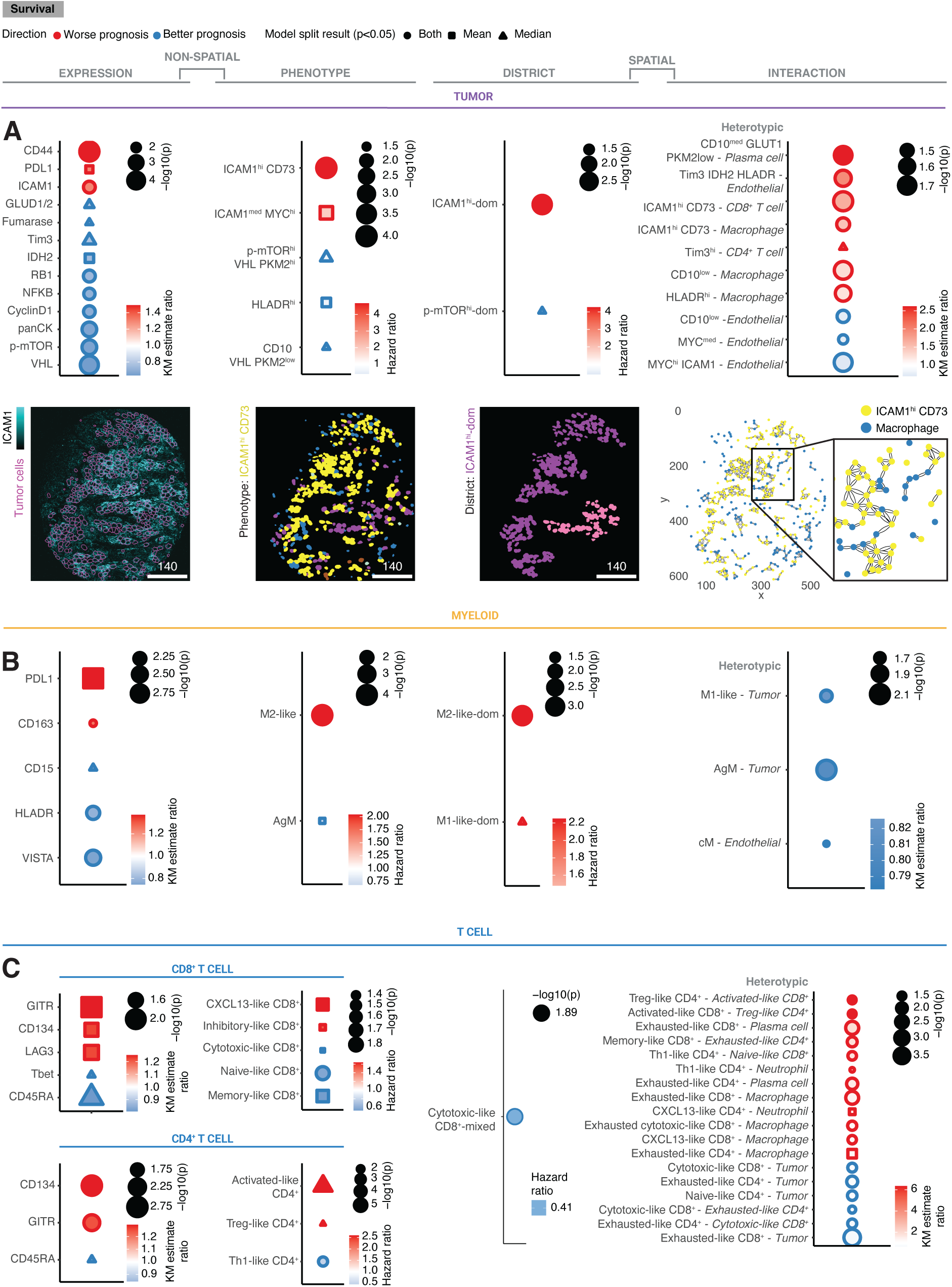
Survival-associated ecosystem features in ccRCC. *See also **Figure S2 and Figure S3**.* ***(A-C)*** Bubble plots of associations of marker expression (left), single-cell phenotypes (center-left), districts (center-right), and cell-to-cell interactions (right) across ***A)*** tumor cells, ***B)*** myeloid cells, and ***C)*** T cells. Bubble color indicates Cox model hazard ratios (phenotype, district) or Kaplan-Meier estimate ratios (expression, interaction). Bubble outline, shape, and size reflect prognostic direction, model results for the indicated model splits, and significance, respectively. Only features with at least one significant association are shown (*P* < 0.05). Representative images of each feature category are shown for tumor features. KM, Kaplan-Meier; AgM, antigen-presenting-like macrophages; cM, classical-like monocytes.

In myeloid cells, high levels of PDL1 and CD163 expression were associated with worse survival, whereas VISTA, HLADR, and CD15 expression were correlated with longer survival **(Figure 2B)**. M2-like macrophages and antigen-presenting-like macrophages were enriched in patients with bad and good survival, respectively **(Figure 2B and Figure S2D)**. Myeloid districts of M2-like and M1-like macrophages were negatively prognostic **(Figure 2B and Figure S2E)**, whereas interactions of antigen-presenting-like macrophages and M1-like cells with tumor cells and of classical-like monocytes with endothelial cells were positively prognostic.

In the T cell compartment, higher levels of GITR and CD134 were associated with worse prognosis, and CD45RA upregulation was associated with better prognosis for both CD8^+^ and CD4^+^ T cells **(Figure 2C)**. For CD8^+^ T cell phenotypes, we found that the presence of naïve-, memory- and cytotoxic-like CD8^+^ T cells were associated with more favorable prognosis, whereas CXCL13-like and inhibitory-like CD8^+^ T cells were indicators of poor survival **(Figure 2C and Figure S2F)**. For CD4^+^ T cell phenotypes, activated and Treg-like CD4^+^ T cells were linked with worse prognosis, and Th1-like CD4^+^ T cells were associated with good prognosis. At the spatial level, districts of cytotoxic-like CD8^+^ T cells mixed with other T cell phenotypes were positively prognostic **(Figure 2C and Figure S2G)**. Among numerous prognostic heterotypic interactions of T cell phenotypes, our analysis revealed that interactions of exhausted and exhausted cytotoxic-like CD8^+^ T cells with macrophages were an indicator of poor prognosis, as previously suggested^9^.

We further investigated the relationships of identified ecosystem features with clinical (ISUP grade, T stage, necrosis) and immunological (PDL1-IMC score) proxies of disease progression. Differential abundances of ecosystem features were observed including higher abundances of MDSC-like phenotypes and related districts in necrotic and PDL1^+^ patients, and more exhausted cytotoxic-like CD8^+^ and CD4^+^ phenotypes and related districts categories associated with disease progression **(Figure S3A, B)**.

In summary, we identified molecular, phenotypic, cell-to-cell interaction, and higher-order spatial single-cell features of tumor and immune cells that are associated with patient survival and disease progression in ccRCC.

### Key ecosystem features define the survival ecotypes of ccRCC

To identify the most important TME properties that drive clinical outcome in ccRCC, we next compared all ecosystem features that were survival- and/or progression-associated in their ability to predict OS in a combined analysis.

We used the statistical machine-learning method *BlockForest*^30^, which has performed well in recent benchmark studies^31,32^. *BlockForest* is an extension of the random survival forest algorithm^33^, in which predictors are grouped into predefined blocks that correspond to different data layers for model training (e.g., ecosystem features and clinical covariates). During tree construction, predictor sampling accounts for the block structure, ensuring that each layer contributes to the model and prevents high-dimensional layers (e.g., ecosystem features) from dominating lower-dimensional but survival-relevant ones (e.g., clinical covariates). To ensure robust cross-comparability, we preprocessed the input feature sets with collinearity reduction, missing value handling and numerical standardization. We used the censored OS time as our model prediction target and evaluated model performance using the Integrated Brier Score and the Concordance-Index (i.e., Uno^34^). The Concordance-Index evaluates discrimination, reflecting the ability of predicted risk scores to correctly rank observed survival times, whereas the Integrated Brier Score assesses overall prediction accuracy over time, incorporating both calibration of predicted survival probabilities and discrimination **(Figure 3A)**. Inclusion of OS times in the prediction framework served as a positive control for model performance **(Figure S4A, B)**.

**Figure 3:**
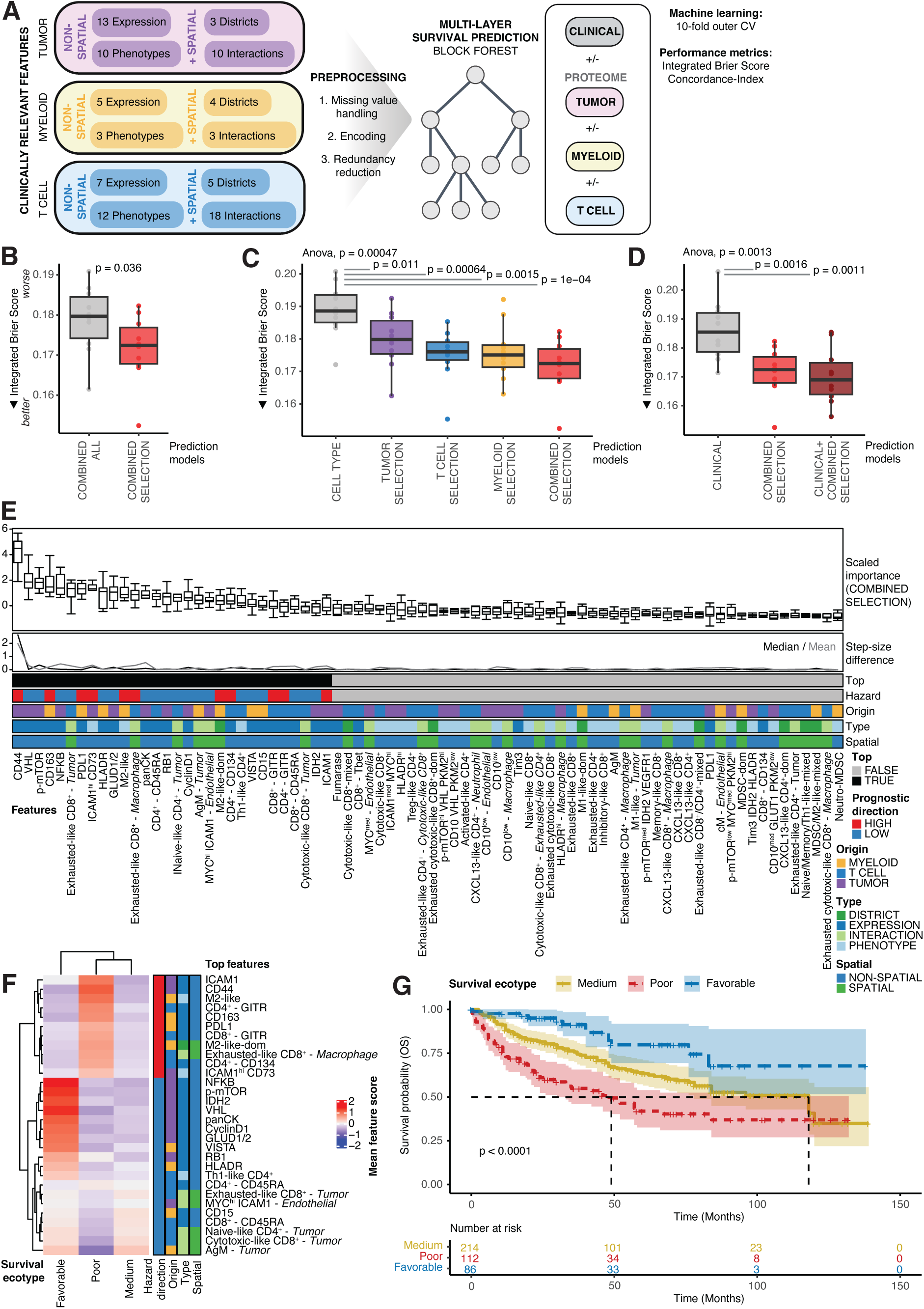
Ecosystem features outperform basic clinical data for survival prediction. *See also **Figure S4***. ***(A)*** Schematic of the workflow for survival prediction (created with BioRender.com). CV, cross-validation. ***(B-D)*** Box plots of Integrated Brier Scores across 10-fold outer cross-validation comparing ***B)*** model with all ecosystem features (COMBINED ALL) versus a model with selected survival-relevant ecosystem features across compartments (COMBINED SELECTION), ***C)*** a model with densities of broad cell types only (CELL TYPE) versus models with selected survival-relevant ecosystem features within and across compartments (TUMOR SELECTION, T CELL SELECTION, MYELOID SELECTION, COMBINED SELECTION), and ***(D)*** a model with basic clinical features (age, gender, T stage) versus a model with selected survival-relevant ecosystem features (COMBINED SELECTION) and a model combining the two (CLINICAL + COMBINED SELECTION) (n=412 patients). Lower Integrated Brier Scores indicate better calibration and discrimination of predicted survival times. Individual points show the test estimate for a single cross-validation run. Box plots show the median, interquartile range (IQR), and whiskers extend to the most extreme values within 1.5×IQR from the quartiles. The *P* values were determined using unpaired one-sided t-tests versus relevant baselines and one-way ANOVA across groups. ***(E)*** Box plot of scaled feature importances across 10-fold outer cross-validation in the COMBINED SELECTION model. Step-size differences between ordered features (median and mean), top features, prognostic directions, feature origin, feature type, and spatial annotations are shown. ***(F)*** Heatmap of mean top 30 feature scores for the *Poor*, *Medium*, and *Favorable* patient survival ecotypes. Prognostic directions, feature origin, feature type, and spatial annotations are shown. ***(G)*** Kaplan-Meier curves of OS for the *Poor*, *Medium*, and *Favorable* patient survival ecotypes. Confidence interval is shown. *P* values were estimated using a log-rank test.

First, we observed that models based only on those ecosystem features identified as associated with OS and/or disease progression performed better than models that used all measured ecosystem features **(Figure 3B and Figure S4C)**, in line with an observed lack of noise resistance of current survival prediction frameworks^31^. Next, we found that compartment-specific models (i.e., those using features from tumor, T cell, or myeloid cells) outperformed a model based simply on densities of clinically relevant broad cell types (e.g, neutrophils and macrophages), which highlights that detailed single-cell and spatial information for these compartments is necessary for better survival prediction **(Figure 3C and Figure S4D-F)**. A combined model including all three compartments performed best. Interestingly, our combined IMC-derived ecosystem feature model also outperformed a model based on clinical features (i.e., age, gender, T stage), but a model combining both clinical and ecosystem information performed best **(Figure 3D and Figure S4G)**.

To identify those ecosystem features that drive performance of the combined IMC model, we analyzed feature importances for survival prediction across model runs **(Figure 3E and Figure S4H)**. Generally, we observed that myeloid- and tumor-related features were more important for prediction of survival than were T cell features, that cell type-specific expression patterns and heterotypic interaction patterns were more informative than phenotypes and districts, and that, overall, our chosen non-spatial metrics outperformed our chosen spatial metrics **(Figure S4H)**. We then defined key survival features as the 30 best performing features, after which step-size differences were minimal **(Figure 3E and Figure S4I)**. Notably, this feature set contained both non-spatial and spatial metrics and features from all cellular compartments. Expression of CD44, a marker of cancer stemness, and VHL, a key tumor suppressor in ccRCC, were the most important predictors overall and for the tumor cell compartment, whereas expression of CD163, a marker of M2 macrophage differentiation, and PDL1, a key immune checkpoint, on myeloid cells and interactions of exhausted-like CD8^+^ T cells with tumor and macrophages were leading predictors in the other compartments.

Subsequently, we performed unsupervised clustering of patients on this set of ecosystem features and identified three distinct outcome groups **(Figure 3F and Figure S4J, K)**: Those in the *Poor* group had high scores for all negatively prognostic features, including expression of ICAM1 and CD44 on tumor cells, the presence of M2-like macrophages and districts, and exhausted-like CD8^+^ T cell interactions with macrophages, all suggestive of a highly immunosuppressive TME state; further, this group of patients was enriched in progressed disease states **(Figure S4L)**. In contrast, patients in the *Favorable* group showed high expression of VHL, NFKB, CyclinD1, and p-mTOR on tumor cells, myeloid cells with upregulated HLADR and VISTA expression, and the presence of Th1-like CD4^+^ cells, pointing towards an immune-activated TME state. The *Medium* group encompassed most patients. This group had higher scores for various immune-to-tumor interactions and the highest endothelial cell density **(Figure S4M)**. OS differed significantly between the groups with *Favorable* and *Poor* patients having the best and worst OS, respectively **(Figure 3G)**. Moreover, the groups provided independent prognostic information for survival beyond that explained by standard clinical features (age, gender, T stage) **(Table S3)**.

Collectively, these analyses showed that single-cell and spatially resolved information on tumor, myeloid, and T cell compartments in ccRCC improved the accuracy of survival prediction over clinical features and allowed the identification of survival ecotypes with differential prognosis.

### Genomic and metabolic alterations are linked with survival ecotypes

The genetic and metabolic make-up of ccRCC is complex^15,16^, and its relation to ecosystem organization and clinical outcome poorly understood. To characterize these aspects of our ecosystem-based survival ecotypes, we analyzed targeted genomic sequencing data with a customized ccRCC specific gene panel in our discovery cohort using published^35^ and newly generated data (n=173 patients with OS information).

The most frequently mutated driver genes included *VHL*, *PBRM1*, *BAP1*, and *SETD2* **(Figure 4A)**, in agreement with previous reports^17,19^. We found strong associations of *TP53* and *BAP1* mutations with decreased survival and progressive disease, respectively **(Figure S5A, B)**, which matches other studies^36,37^. Genomic mutation information alone performed worse for survival prediction than IMC-derived ecosystem features but slightly improved the prediction in combination with ecosystem features **(Figure S5C)**. Survival ecotypes remained prognostic in the subset of patients with available sequencing data **(Figure S5D).** Further, the patients in the *Poor* group harbored significantly more *BAP1* mutations, those in the *Favorable* group lacked *BAP1* and *PBRM1* mutations, and in the *Medium* group there was a tendency toward *PBRM1* mutations **(Figure 4B)**. Mutational burden was not different between ecotypes and was not prognostic **(Figure S5E, F)**.

**Figure 4:**
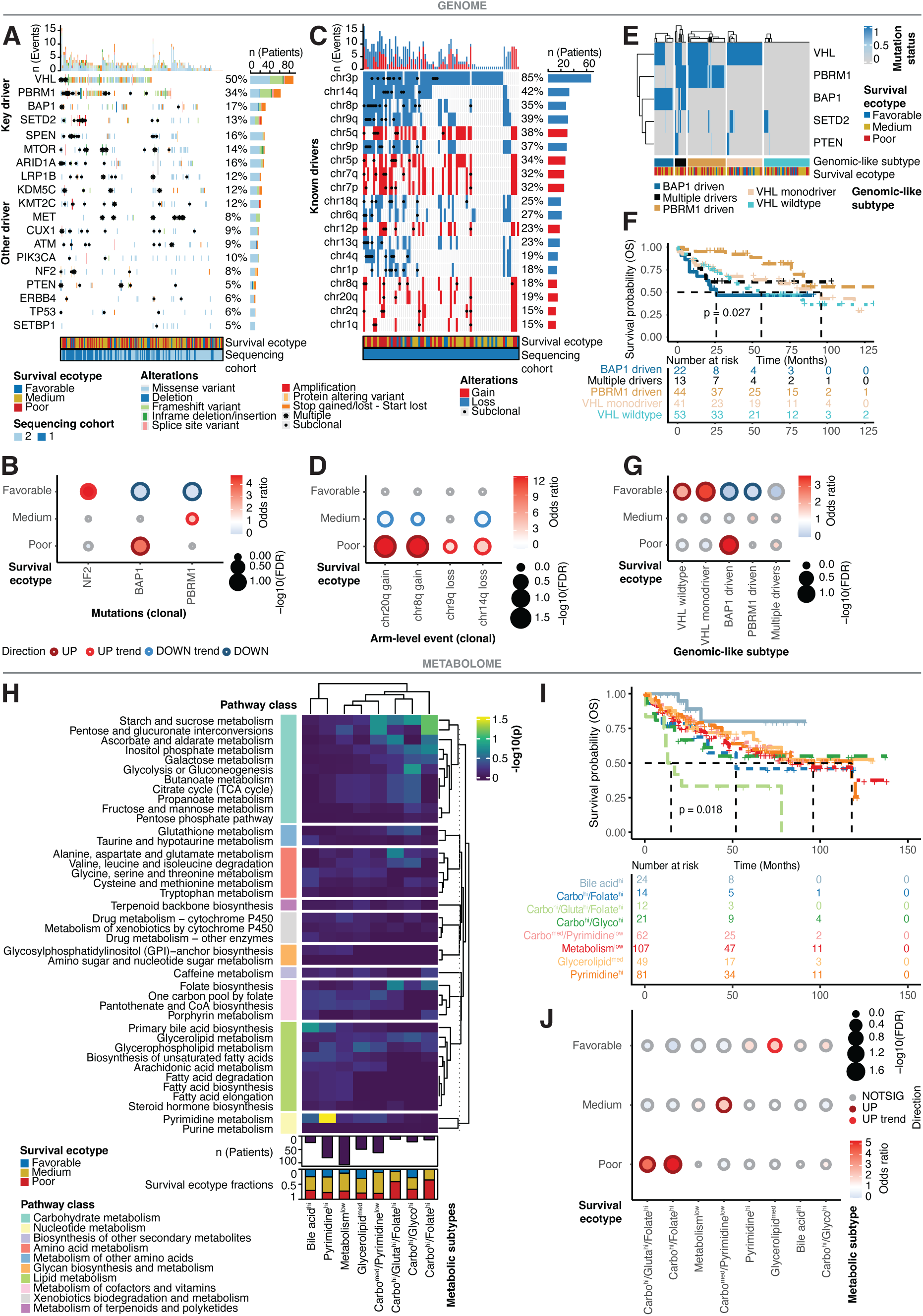
Survival ecotypes have distinct genomic and metabolic features. ***See also*** Figure S5-S7. ***(A)*** Oncoprint of driver mutations (n=173 patients with OS information). Alterations per gene and patient are displayed (top and right). Survival ecotypes and sequencing cohort are annotated (bottom). ***(B)*** Bubble plot of associations of survival ecotypes with mutations (clonal). Bubble color, outline, and size reflect odds ratio from Fisher exact testing, association direction, and significance, respectively. Only mutations with at least one significant association are shown (FDR < 0.1; trends at 0.05). ***(C)*** Oncoprint of driver CNV arm-level events (n=79 patients with OS information). Alterations per arm and patient are displayed (top and right). Survival ecotypes and sequencing cohort are annotated (bottom). ***(D)*** Bubble plot of associations of survival ecotypes with arm-level events (clonal). Bubble color, outline, and size reflect odds ratio from Fisher exact testing, association direction, and significance, respectively. Only events with at least one significant association are shown (FDR < 0.1; trends *at P* < 0.05). ***(E)*** Heatmap of mutation status of selected genes per patient, split by genomic-like subtype. Survival ecotypes are annotated. ***(F)*** Kaplan-Meier curves of OS for genomic-like subtypes. *P* values were estimated using a log-rank test. ***(G)*** Bubble plot of associations of survival ecotypes with genomic-like subtypes. Bubble color, outline, and size reflect odds ratio from Fisher exact testing, association direction, and significance (FDR < 0.1; trends at *P* < 0.05), respectively. ***(H)*** Heatmap of pathway enrichment results for metabolic subtypes defined based on MALDI-MSI data (n=443 patients). Rows are split by pathway class. Number of patients and survival ecotype fractions per group are shown. ***(I)*** Kaplan-Meier curves of OS for metabolic subtypes (n=370 patients with OS information). *P* values were estimated using a log-rank test. ***(J)*** Bubble plot of associations of survival ecotypes with metabolic subtypes. Bubble color, outline, and size reflect odds ratio from Fisher exact testing, association direction, and significance (FDR < 0.1; trends at *P* < 0.05), respectively.

Chromosome arm level events were analyzed in a subset of the cohort with matched normal samples (n=79 patients with OS information). We identified chromosome 3p loss as the most frequent alteration in ccRCC, followed by chromosome 14q/8p/9q losses and 5q gain **(Figure 4C)**, in line with other cohort studies^17,19^. Several of these events were linked to survival and disease progression, including chr9p/14q losses **(Figure S5G, H)**, as described previously^38^. Survival prediction with these arm-level events showed no improvement over ecosystem features **(Figure S5I)**, but survival ecotypes were prognostic in this patient subset **(Figure S5J)**. The *Poor* subset of patients was enriched with chromosome 20q/8q gains and 9q/14q losses, which includes the proto-oncogene *cMYC* on 8q and the tumor suppressor gene *HIF1A* on 14q^39^. These patients had an overall higher arm-level copy number variation (CNV) burden, which was also linked to worse survival **(Figure 4D and Figure S5K, L)**.

Given findings from the TRACERx Renal study^17^, we assessed the co-occurrence patterns of *VHL*, *PBRM1*, *BAP1*, *SETD2*, and *PTEN* mutations and assigned genomic-like subtypes to each patient similar to the original study **(Figure 4E)**. These genomic-like subtypes differed in OS, with *PBRM1*-driven cases having the best survival and *BAP1*-driven cases and cases with multiple drivers having the worst survival **(Figure 4F)**, which is comparable to the original study^17^. The *Favorable* survival ecotype was enriched in subjects with wild-type *VHL* and *VHL* monodriver subtypes and absent in *BAP1*-driven and *PBRM1*-driven cases **(Figure 4G)**. The *Poor* survival ecotype was enriched in *BAP1*-driven subtype patients, in line with the mutation data.

We also analyzed tumor-region specific metabolic profiles from MALDI-MSI data^40^ in our discovery cohort (n=370 patients with OS information). Several analyte species with different m/z values were associated with survival and disease progression (**Figure S6A-D)**, but use of these data did not improve survival prediction over that achieved by the IMC-derived features, either alone or in combination **(Figure S6E)**. We classified patients into eight metabolic subtypes based on clustering of the MALDI-MSI data, pathway enrichment analysis of the clusters, and merging of these clusters based on similarity of their enriched pathways **(Figure 4H and Figure S6F-H)**. The Carbo^hi^/Glutamate^hi^/Folate^hi^ subtype had the worst survival; the *Poor* survival ecotype was enriched with this subtype and with the Carbo^hi^/Folate^hi^ subtype **(Figure 4I, J)**. The *Medium* subtype was enriched in the Carbo^med^/Pyrimidine^low^ metabolic subtype, whereas the *Favorable* subtype had a tendency toward enrichment in a metabolic subtype with lower overall pathway scores.

These data show that our ecosystem-derived survival ecotypes of ccRCC have distinct genomic and metabolic attributes, suggesting that there is a link between TME organization and these properties.

### Survival ecotypes correlate with spatial CD8^+^ T cell architecture

In ccRCC, increased CD8^+^ T cell presence in the tumor is paradoxically associated with worse prognosis^41,42^. We therefore analyzed whether the spatial location of CD8^+^ T cells differs depending on phenotypic profiles and assessed the relationship of spatial locations of these phenotypes to our survival ecotypes. We assigned each CD8^+^ T cell to one of three spatial patterns: Infiltrating, Bystanding, or Ignored **(Figure S7A)**. Assignment was based on the number of direct interactions to tumor cells as described in our previous study^43^. Infiltrating (≥ 3 tumor cell interactions) and Bystanding (1-2 tumor cell interactions) cells were more often cytotoxic- or exhausted-like, whereas Ignored cells (0 tumor cell interactions) were enriched in exhausted cytotoxic-like and naïve-like CD8^+^ **(Figure S7B)**. Based on the ratio and counts of these spatial CD8^+^ T cell patterns, we assigned each patient to one of three spatial CD8^+^ T cell groups: *Inflamed*, *Excluded*, or *Cold* **(Figure S7C)**. Patients in the *Excluded* group had lower survival probabilities than the other groups, and this group was associated with measures of clinical disease progression **(Figure S7D, E)**. In contrast, stratification based on solely CD8^+^ T cell counts per patient was not prognostic **(Figure S7F)**. The *Poor* and *Favorable* survival ecotypes were strongly correlated with *Excluded* and *Inflamed* spatial CD8^+^ groups, respectively **(Figure S7G)**.

These data highlight that the spatial location of CD8^+^ T cells, rather than their presence alone, is important for patient prognosis and that this information is encoded in the survival ecotypes.

### Morphology contains survival ecotype information and enables gene signature extraction

To assess the potential of our survival ecotypes for clinical translation, we tested whether these ecotypes could be predicted from standard pathology H&E images. The H&E images for tumors from our cohort were from consecutive sections to those imaged by IMC. To analyze the images, we used a deep-learning framework based on UNI^44^, a self-supervised “foundation model” for H&E feature extraction, and CLAM^45^, an attention-based, multiple-instance learning method for classification **(Figure 5A and Figure S8A)**. We found that the morphology information in the H&E images, extracted by UNI and classified with CLAM, allowed survival ecotype prediction in the discovery cohort with overall high accuracy (mean weighted ROC-AUC score: 0.84; mean weighted F1 score: 0.69) and with better performance for the *Poor* and *Favorable* subtypes than for the *Medium* subtype **(Figure 5B, C and Figure S8B)**. Moreover, several clusters of UNI-extracted patch-level embeddings were associated with survival and disease progression **(Figure S8C-F)**, yet they did not provide better survival predictions over IMC-derived features, whether considered alone or in combination **(Figure S8G)**.

**Figure 5:**
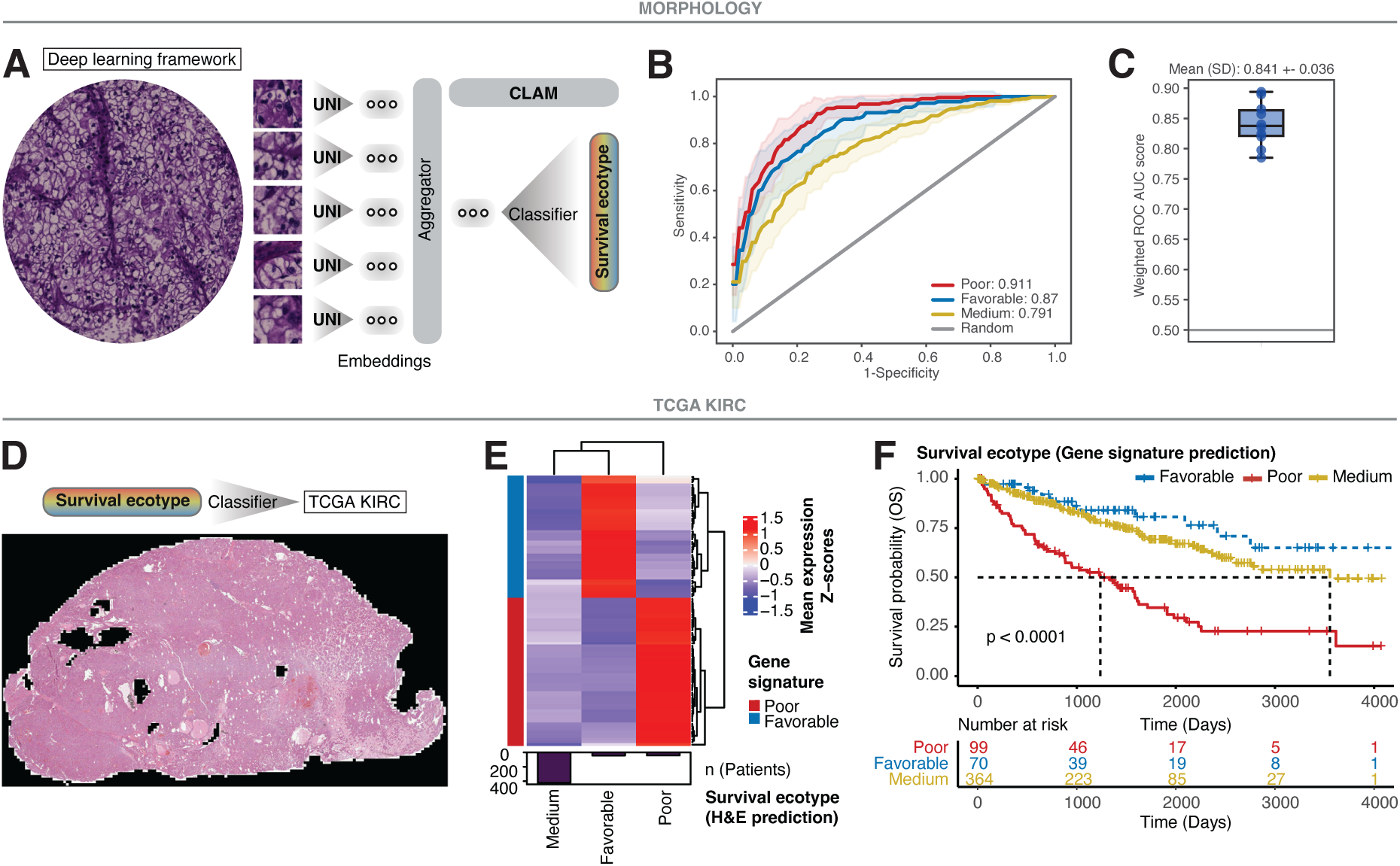
ccRCC survival ecotypes can be predicted from standard pathology images. *See also **Figure S8 and Figure S9***. ***(A)*** Deep learning-based workflow for classification of survival ecotypes from H&E images (created with BioRender.com). ***(B)*** Mean receiver operating characteristic (ROC) test curves for each survival ecotype across 10-fold Monte Carlo cross-validation with confidence intervals (±SD). Random chance diagonal and mean area under the curve (AUC) per ecotype are displayed. ***(C)*** Box plot of weighted ROC AUC test scores across 10-fold Monte Carlo cross-validation for all survival ecotypes. Mean and SD are indicated. Box plots show the median, IQR, and whiskers extend to the most extreme values within 1.5×IQR from the quartiles. ROC, receiver-operating-characteristic; AUC, area under curve. ***(D)*** Representative H&E image from the TCGA KIRC cohort^19^ for survival ecotype prediction from trained classifier. ***(E)*** Heatmap of mean expression Z-scores for all genes in the gene signatures of the *Poor* (n=57 genes) and *Favorable* (n=47 genes) H&E-predicted survival ecotypes in the TCGA KIRC cohort (n=510 patients with H&E data and OS information). Number of patients per group are shown. ***(F)*** Kaplan-Meier curves of OS for gene-signature-predicted survival ecotypes in the TCGA KIRC cohort (n=533 patients with H&E data and OS information). *P* values were estimated using a log-rank test.

Next, we applied the trained classifier to whole-slide H&E images from the TCGA Kidney Renal Clear Cell Carcinoma (KIRC) dataset (n=510 patients with H&E data and OS information)^19^ **(Figure 5D)**, and observed that H&E-predicted survival ecotypes followed similar survival trends as in the discovery cohort, but with less pronounced differences between *Favorable* and *Medium* subtypes **(Figure S8H)**. Since transcriptomic data are available for the TCGA KIRC cohort, we analyzed gene expression patterns in the H&E-predicted survival ecotypes. Gene set enrichment analysis of differentially expressed genes demonstrated upregulation of ion homeostasis processes in the *Favorable* subtype, suggesting more normal-like kidney function, vascular programs in the *Medium* subtype, and cell cycle-related processes in the *Poor* subtype, suggesting genetic instability, which we confirmed with a chromosomal instability signature^46^ **(Figure S8I, J)**. Based on the differentially expressed genes, we then derived gene signatures for the *Poor* (n=57 genes) and *Favorable* (n=47 genes) subtypes with the goal of using these signatures to predict the survival ecotypes in other transcriptomic datasets **(Figure 5E, Table S4)**. For TCGA KIRC subjects (n=533 patients with OS information), gene-signature-predicted survival ecotypes, derived using a nearest template prediction algorithm, had significantly diverging OS probabilities with the *Poor* and *Favorable* groups having the poorest and best survival, respectively **(Figure 5F and Figure S9A)**. This was expected given the similarity of gene-signature-predicted ecotypes to the H&E-predicted ones **(Figure S9B)**. As in the discovery cohort, the gene-signature-predicted ecotypes provided independent prognostic information for survival beyond that explained by gender and T stage **(Table S3)**.

We then tested whether the gene-signature-predicted ecotypes in the TCGA KIRC cohort had comparable molecular attributes as those in our discovery cohort. As in our discovery cohort, for the TCGA KIRC cohort, *ICAM1* and *CD44* expression was highest in the *Poor* group, and *VHL*, *NFKB1*, and *MTOR* levels were elevated in the *Favorable* group **(Figure S9C)**. Deconvolution of the gene expression data revealed the presence of various immune cell types within TCGA KIRC tumors, including M2 macrophages and CD8^+^ T cells in the *Poor* ecotype, CD4^+^ T cells in *Favorable* samples, and endothelial cells in *Medium* patients, in agreement with findings from the discovery cohort **(Figure S9D)**. Also as found in the discovery cohort, in the TCGA KIRC subjects, *BAP1* mutations were enriched in the *Poor* ecotype, and no trends were detected for *PBRM1* mutations **(Figure S9E)**. Moreover, suggestive of a plastic state, *Poor* patients had the highest expression of two recently proposed gene signatures – a genomic dedifferentiation signature^47^ and a de-clear cell differentiated ccRCC (IM4) signature^26^ **(Figure S9F)**. Finally, we tested our defined gene signatures to identify ecotypes in the independent CPTAC patient cohort (n=212 patients)^23^ and confirmed both survival trends and gene expression patterns of key genes in this cohort **(Figure S9G-I)**.

Taken together, these deep-learning based analyses show that information on ccRCC survival ecotypes is captured in standard pathology H&E images, opening a promising avenue for clinical translation.

### Treatment responses of survival ecotypes differ

To corroborate the prognostic impact of the survival ecotypes and identify potential treatment targets, we next used our deep-learning based gene signatures to predict the ecotypes in bulk transcriptomic data for patients from three large clinical trials of RCC: JAVELIN 101^4^, CheckMate^48^, and IMmotion 151^24^.

The JAVELIN 101 trial (n=733 patients) compared a first-line combination of immunotherapy (a PDL1 inhibitor, avelumab) plus VEGFR inhibition (axitinib) to treatment with a tyrosine kinase inhibitor (sunitinib) alone for untreated advanced RCC with a clear-cell component. In this cohort, we found that the OS differences between the survival ecotypes were recapitulated in the full cohort, driven by patients in the combination arm **(Figure 6A and Figure S10A, B)**. Progression-free survival (PFS) showed similar trends with survival differences across the full cohort and treatment arms. Interestingly, *Medium* patients significantly benefited from the combination treatment in terms of OS and PFS, and showed higher treatment responses in this arm, with the *Poor* group also displaying a significant PFS and treatment response effect **(Figure 6B, C)**. Notably, the ecotypes provided independent prognostic information for OS beyond the IMDC score, a prognostic model based on clinical parameters that is used for clinical patient stratification^49,50^ **(Table S5)**. Expression of key genes followed trends we had observed in the discovery and TCGA KIRC cohorts, namely expression of *ICAM1* and *CD44* in *Poor* patients, suggestive of migration and stem-like traits, and of *VHL*, *CCND1*, *NFKB1*, and *MTOR* in *Favorable* patients, suggestive of an immune-activated and VHL-intact state **(Figure S10C)**.

**Figure 6:**
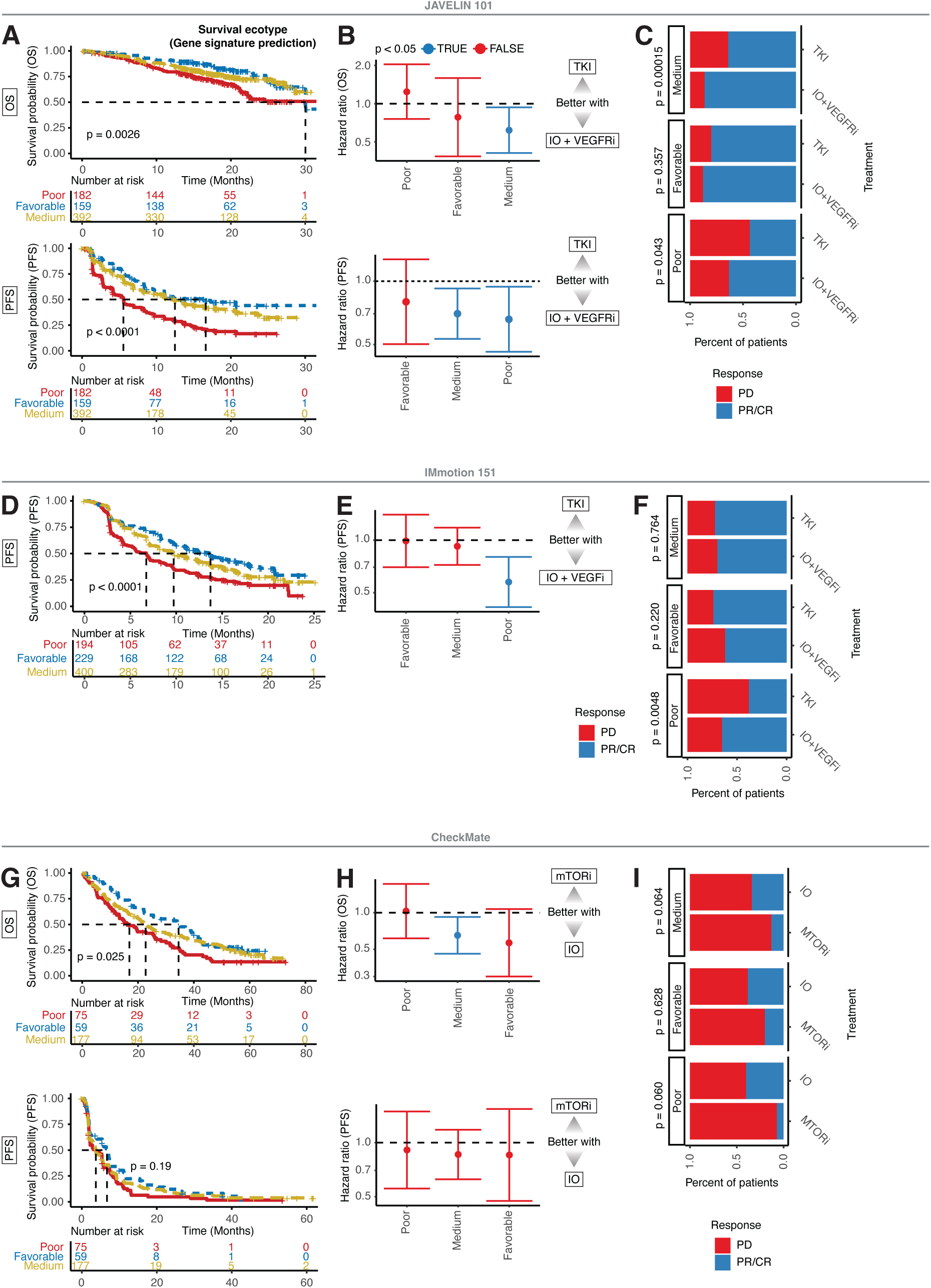
Immunotherapy response and survival differs between ecotypes in clinical trial datasets. ***See also*** *Figure S10*. ***(A)*** Kaplan-Meier curves of OS (top) and PFS (bottom) for gene-signature-predicted survival ecotypes in the JAVELIN 101 cohort^4^ (n=733 patients). *P* values were estimated using a log-rank test. ***(B)*** Hazard ratios of OS (top) and PFS (bottom) estimated using a Cox proportional hazards model (Wald test) comparing treatment arms of JAVELIN 101 across gene-signature-predicted survival ecotypes. Error bars show the 95% confidence intervals. IO, immunotherapy; TKI, tyrosine kinase inhibitor ***(C)*** Stacked bar plots of treatment responses in the two treatment arms of JAVELIN 101, shown for all gene-signature-predicted survival ecotypes. *P* values were determined by Fisher exact testing. ***(D)*** Kaplan-Meier curves of PFS for gene-signature-predicted survival ecotypes in the IMmotion 151 cohort^24^ (n=823 patients). *P* values were estimated using a log-rank test. ***(E)*** Hazard ratios of PFS estimated using a Cox proportional hazards model (Wald test) comparing treatment arms of IMmotion 151 across gene-signature-predicted survival ecotypes. Error bars show the 95% confidence intervals. ***(F)*** Stacked bar plots of treatment responses in the two treatment arms of IMmotion 151, shown for all gene-signature-predicted survival ecotypes. *P* values were determined by Fisher exact testing. ***(G)*** Kaplan-Meier curves of OS (top) and PFS (bottom) for gene-signature-predicted survival ecotypes in the CheckMate cohort^48^ (n=311 patients). *P* values were estimated using a log-rank test. ***(H)*** Hazard ratios of OS (top) and PFS (bottom) estimated using a Cox proportional hazards model (Wald test) comparing treatment arms of CheckMate across gene-signature-predicted survival ecotypes. Error bars show the 95% confidence intervals. ***(I)*** Stacked bar plots of treatment responses in the two treatment arms of CheckMate, shown for all gene-signature-predicted survival ecotypes. *P* values were determined by Fisher exact testing.

The IMmotion151 trial (n=823 patients) monitored PFS and treatment response in patients with untreated advanced RCC with clear-cell/sarcomatoid characteristics treated with a combination of a PDL1 inhibitor (atezolizumab) and a VEGF-A inhibitor (bevacizumab) or only with the tyrosine kinase inhibitor sunitinib. PFS was again significantly different between survival ecotypes, both overall and in the sunitinib arm **(Figure 6D and Figure S10D, E)**. *Poor* patients in the combination arm had improved PFS and better treatment responses than those in the sunitinib treatment arm **(Figure 6E, F)**. *ICAM1* and *CD44* were predominantly expressed in the tumors of the *Poor* group, and *NFKB1* and *CCND1* were enriched in the tumors of *Favorable* patients **(Figure S10F)**. Moreover, survival ecotype information was independently prognostic beyond the IMDC score **(Table S5)**.

The CheckMate trial (n=311 patients) included previously treated patients with advanced RCC with a clear cell component. One group was given the PD1 inhibitor nivolumab and the other the mTOR inhibitor everolimus. Our analysis confirmed OS differences for the survival ecotypes both overall and in the nivolumab arm **(Figure 6G and Figure S10G, H)**. *Medium* patients survived significantly longer overall with PD1 inhibitor treatment than with mTOR inhibitor treatment **(Figure 6H)**, and both *Poor* and *Medium* patients showed trends for better responses with the PD1 inhibitor treatment **(Figure 6I)**. PFS analysis showed no significant differences between ecotypes, but gene expression patterns in this cohort were comparable to those observed for subjects from the IMmotion151 trial **(Figure S10I)**, and ecotype encoded OS information was again independent of the IMDC score **(Table S5).**

In summary, these findings demonstrate the clinical relevance of our proposed survival ecotypes, especially in the current era of immunotherapy for ccRCC.

## DISCUSSION

In management of ccRCC, oncologists do not consider molecular features, and survival benefits of existing treatments are variable^7^. Patient survival is influenced by the multifaceted interplay of diverse cell types in the TME, but the ccRCC ecosystem had not been comprehensively characterized spatially at single-cell resolution. In this multi-omic study, we made use of highly multiplexed imaging data of tumor, myeloid, and T cell architectures within ccRCC tumors to identify survival-relevant ecosystem features that encompassed both spatial and non-spatial properties of the TME. This feature set formed the basis for delineating three survival ecotypes of ccRCC that we validated across independent patient cohorts from multiple international sites.

Patient tumors of the *Poor* survival ecotype were characterized by high levels of ICAM1 and CD44 protein expression on tumor cells and a previously described immune dysfunction circuit of M2-like macrophages and T cell exhaustion and exclusion^9–11^. ICAM1, a cell surface glycoprotein that regulates cellular interactions, was previously reported to be associated with metastatic tumor progression and migration, and poor prognosis of patients with ccRCC^51–53^. ICAM1 is mechanistically connected to T cell suppression via tumor-derived exosomes^54^, and it associates directly with CD44, a canonical cancer stem cell marker^55,56^, to induce trans-endothelial leukocyte migration^57^. That the tumor cells of the *Poor* ecotype are stem-like was further supported by enrichment in two recently proposed gene signatures of dedifferentiation^26,47^. We also observed association of *Poor* patients with BAP1 mutation, higher CNV burden and genetic instability, which may be linked given that loss of BAP1 function compromises DNA repair and in turn genomic instability is linked to tumor plasticity and inflammation^58,59^. These findings suggest a convergence of genetic instability, tumor plasticity and migration, and immune evasion to yield a bad prognosis for the *Poor* survival ecotype. Our analysis of clinical trials, JAVELIN 101^4^, CheckMate^48^, and IMmotion 151^24^, suggested that the *Poor* group patients respond to immunotherapy approaches but that this response does not result in sustained OS benefits. This highlights the need for novel treatment approaches. Based on our findings, antibody-based targeting of ICAM1, which has shown promise in other cancer types either alone^60^, in combination with immunotherapy^61^, in form of antibody-drug conjugates^62,63^ and as CAR T cells^64^ (NCT04420754), could be considered for treatment of patients of the *Poor* ecotype.

On the other end of the prognostic spectrum, tumors of the *Favorable* survival ecotype showed higher protein expression of VHL and associations with VHL monodriver/wild-type genomic-like subtypes, suggesting that the tumor suppressing function of VHL via HIF-dependent or independent pathways are at least partly retained in these tumors^15,65^. It was previously reported that in patients with ccRCC, higher expression of VHL is correlated with better survival and increased immunogenicity^66^. Our data supports this as we observed NFKB tumor expression and the enrichment of antigen-presenting myeloid cells, Th1-like CD4^+^ T cells, and direct interactions of CD8^+^ T cells with tumor cells in patients of the *Favorable* ecotype. Elevated CyclinD1 expression was also detected in these tumors; CyclinD1 is a downstream effector of the VHL-HIF signaling axis that was previously associated with favorable outcome for RCC^67^. CyclinD1–specific CD8^+^ T cell responses have been observed in subjects with ccRCC^68^. Moreover, a transcriptional program resembling that of normal kidney epithelial cells was observed in patients of the *Favorable* ecotype, suggesting that epithelial differentiation is maintained. Together, these characteristics point towards an immune-activated, VHL-intact, and normal-like tumor cell state in the *Favorable* survival ecotype. Treatment-wise, we observed high overall response rates for these patients to all treatments evaluated in the JAVELIN 101^4^, CheckMate^48^, and IMmotion 151^24^ trials.

We observed significant OS benefits for immunotherapy approaches for patients with tumors of the *Medium* survival ecotype, the ecotype of most patients in all cohorts evaluated. This group has more heterogenous tumors than those of patients in *Poor* and *Favorable* groups. Few conserved and pronounced TME features were identified, but *Medium* tumors had a vascularized ecosystem with trends for *PBRM1* mutations, consistent with previously observed responses to checkpoint therapy for patients with *PBRM1* mutations and an immune desert and endothelial cell rich phenotype previously described^22,25,48,69^.

This study has limitations. First, for our retrospective discovery cohort, follow-up treatment information was missing, which may have confounded the survival analysis due to unmeasured treatment effects. However, treatment information was available for three validation cohorts (JAVELIN 101, CheckMate, and IMmotion 151). Second, our intra-tumoral heterogeneity assessment of the discovery cohort data was limited since we analyzed relatively small pathologist-chosen tumor areas with IMC; this strategy was used because it allowed us to analyze many patients. For most patients, at least two IMC images per panel were analyzed. Third, genomic and metabolomics data were analyzed from bulk samples rather than at the single cell level, although genomic data for two biopsies were available for around half of the patients.

Although future prognostic and predictive validation will be needed to assess the utility of our proposed survival ecotypes in clinical practice, our ecosystem-based stratification approach outperformed standard clinical data in predicting patient survival for the more than 2500 patients evaluated. We showed that these ecotypes are identifiable from standard H&E images before treatment, thus paving the way for incorporating this classification system into personalized patient stratification strategies. In conclusion, the three survival ecotypes of ccRCC proposed here are based on spatial single-cell features and have potential as future biomarkers for improved patient stratification.

## METHODS

### Discovery cohort

The patient samples and associated clinical information for the discovery cohort were obtained from the Department of Pathology and Molecular Pathology of the University Hospital Zurich. Expert pathologists (H.M.) designed and constructed two tissue microarrays (TMAs), TMA20 and TMA224, from pre-treatment tumor samples from patients enrolled between 1993 and 2013^70^. The TMAs contained 0.6-mm diameter cores from tumor regions with at least 70% tumor cells, with one to six cores per patient after quality control. The study was approved by the Zurich Ethical Commission under Ethical approval number BASEC: 2019-01959.

### Imaging mass cytometry

#### Antibody panels

Three antibody panels were designed to separately analyze tumor cells, T cells, and myeloid cells of ccRCC in detail. Antibodies targeted 85 unique compartment-specific markers, cellular state/immunoregulatory proteins, and general cell type markers. More detailed information on the panels can be found in **Table S1**.

#### Tissue preparation and staining

Tissue samples were formalin-fixed and paraffin-embedded (FFPE) at the University Hospital Zurich. Tissue sections of 2 µm thickness were stained with the antibody panels as previously described^71^ and deparaffinized in xylene three times for 10 min each wash before being rehydrated in a graded alcohol series (ethanol:deionized water, 2x 100:0, for 5 min; 2x 96:4, 90:10, 80:20, 70:30, 3 min each). Antigen retrieval was conducted with Tris-EDTA (pH 9) buffer at 95 °C in a NxGen decloaking chamber (Biocare Medical) for 30 min. Following cooling for 20 min, slides were blocked with 3% BSA 0.1% Tween in 150 mM NaCl, 50 mM Tris-HCl, pH 7.6 (TBS) for 1 h. Samples were first stained and incubated with untagged rabbit anti-CD4 antibody overnight at 4 °C. Samples were then washed in TBS before incubation with metal-tagged anti-rabbit secondary antibody (2 h at room temperature) to increase CD4 signal. After washing with TBS again, samples were incubated with the metal-tagged antibody panel overnight at 4 °C. Following incubation, samples were stained with 0.5 µM Cell-ID Intercalator-Ir (Fluidigm, 201192B) for detection of DNA. After 7 min, slides were rinsed with TBS three times for 10 min each wash and then briefly rinsed in water and air dried.

#### Image acquisition

IMC images were acquired using a Hyperion Imaging System (Standard BioTools). The largest square area of each core was laser-ablated at 400 Hz. Cores were randomly selected for acquisition, and data on the two TMAs were acquired in one continuous run. When acquisitions were unexpectedly interrupted, the remaining portion of the core was acquired as a separate image, leading to a small number of cores divided into two images. After quality control, a total of 2924 images were selected for further analysis.

#### Data processing

Raw data processing was performed using *steinbock* (v.0.16.1), following the IMC data analysis workflow^72^ available at https://github.com/BodenmillerGroup/IMCDataAnalysis. After extracting multi-channel images from raw IMC data, which includes hot-pixel filtering, cell segmentation was performed with the pre-trained deep-learning based Cellpose model^73^ (v.2.2.2, Python 3.9.17) using the same markers across panels (Nucleus: Ir191, Ir193, H3; Cytoplasm: CA9, panCK, SMA, CD8a, CD68). We corrected pixel-artifacts across images using a customized in-house script. For each image, we then obtained mean pixel intensities per cell and marker, morphological features (e.g., area, eccentricity), and neighborhood information (based on a spatial cell graph constructed by pixel expansion). We estimated tissue areas per core using a pipeline combining *ilastik* (v.1.4.0)^74^ and *CellProfiler* (v.4.0.7)^75^.

All downstream analyses of IMC and other data types described below were performed in R (v.4.3.2) on Ubuntu if not stated otherwise. Single-cell data and images were read into R using the *imcRtools* (v.1.8.0) and *cytomapper* (v.1.14.0)^76^ R/Bioconductor packages, respectively. We used the *SingleCellExperiment* object (v.1.24.0)^77^ as a data container for downstream analysis. Mean pixel intensities per marker and cell were arsinh-transformed using a cofactor of 1 and are referred to as expression values. Spillover compensation was performed with the *CATALYST* R/Bioconductor package (v.1.26.0) as previously described^78^. To ensure robust downstream data analysis, we visually inspected images using the *cytoviewer* R/Bioconductor package (v.1.2.0)^79^ and excluded those with low total marker expression across channels, low total number of cells, small tissue area, or small fraction of tissue area covered by cells. Furthermore, cells with low nuclear expression (sum of expression values across DNA channels H3, ^191^Ir, and ^193^Ir below 1) were excluded. These quality control steps yielded 494 patients, 2924 images, and a total of 3,211,104 cells for downstream IMC data analysis.

#### Image visualization

Pixel and single-cell level IMC images were generated using the *cytoviewer* R/Bioconductor package (v.1.2.0)^79^.

#### Cell- and phenotype prediction

In addition to unsupervised clustering (described below), we used two separate methods to support 1) tumor and non-tumor cell separation, 2) identification of broad TME cell types and 3) fine-grained classification of myeloid and T cell phenotypes. First, we used the Python-based *astir* (v.0.1.4, Python 3.9.7) tool, a probabilistic deep recognition neural network inference-based modeling framework built to assign known cell types based on key marker information^29^. Second, we employed Gaussian mixture modelling (GMM) using the *mclust* (v.6.0.1) R package to classify cells as positive or negative for key markers. Detailed information on used marker sets for each task and probability/uncertainty thresholds can be found in **Table S2**.

#### Tumor and broad cell-type classification

For tumor and non-tumor cell separation, we applied self-organizing map-based clustering on expression values from relevant shared markers across panels (CA9, panCK, CD45, CD31/vWF, SMA, CD3, CD4, CD8a, CD38, MPO, CD11c, CD20, CD68) using the *CATALYST* R/Bioconductor package (v.1.26.0). We evaluated cluster stability using different metrics, including silhouette width and root mean-squared deviation, and selected 24, 16, and 23 clusters for analysis in the tumor, myeloid, and T cell panels, respectively. Next, we assigned one of three cluster categories (tumor, mixed, non-tumor) to each cluster based on the fraction of non-tumor cells predicted by *astir* (tumor panel: tumor ≥50%; mixed, 15-50%; non-tumor, <15% - Myeloid/T cell panel: tumor ≥50%; mixed, 10-50%; non-tumor, <10%). Lastly, we annotated cells in mixed clusters using the GMM predictions to either the tumor or non-tumor compartment.

For classification of the non-tumor cells, we used graph-based clustering (Jaccard index-based weights, Louvain algorithm for community detection) on expression values from shared key markers across panels (CD45, CD31/vWF, SMA, CD3, CD4, CD8a, CD38, MPO, CD11c, CD20, CD68) with different numbers of nearest neighbors (*k* = 30, 50, 70, 90, 110, 130) using a modified R implementation of the *PhenoGraph* algorithm (*Rphenoannoy*; v.0.1.0) (https://github.com/stuchly/Rphenoannoy). Cluster stability assessment was performed, and we selected *k* values of 110, 90, and 110 for analyses of the tumor, myeloid, and T cell panels, respectively. Next, we assigned broad TME cell types in two steps: First, all clusters with canonical marker expression and high *astir* and GMM prediction fractions for a given cell type were assigned to this cell type. Cells from remaining clusters were assigned first based on GMM predictions and afterwards, if still unassigned, based on *astir* predictions (tumor and myeloid panels) or re-clustered using self-organizing map-based clustering and then assigned based on interpretation of marker expression (T cell panel).

#### Phenotype classification

For phenotype classification, we pooled relevant broad cell types for each panel (tumor panel: tumor cells; myeloid panel: macrophages and neutrophils; T cell panel: CD4^+^ T cells, CD8^+^ T cells) for clustering with compartment-specific markers (tumor panel: GPX4, HLADR, NFKB, CyclinD1, HIF1a, p53, cMYC, ICAM1, PDK4, CD73, BRG1, PKM2, PDL1, CD10, IDH2, HIF2a, AuroraKinaseA, Tim3, EGFR, p-S6, VEGFR1, VHL, RB1, p-mTOR, GLUT1, Fumarase, GLUD1/2; myeloid panel: CD68, CD11c, MPO, CD16, HLADR, CD163, CD11b, MMP9, CCR2, Arginase, CD33, CD209, CD206, CD15; T cell panel: CD4^+^ T cell: CD45RA, LAG-3, Tim3, PD1, NKG2A, CXCL13, EOMES, Tbet, FOXP3, GATA3, GITR, CD134, CD278; CD8^+^: CD45RA, LAG-3, Tim3, PD1, NKG2A, CXCL13, EOMES, Tbet, GITR, CD134, CD278, GranB, GranK) using the *PhenoGraph* algorithm across different numbers of neighbors (*k* = 30, 50, 70, 90, 110, 130). Cluster stability assessment was performed, and we selected *k* values of 130, 90, and 130 for CD8^+^ T cells in the tumor, myeloid, and T cell panels, respectively and a *k* value of 70 for CD4^+^ T cells in all panels. Next, we assigned fine-grained phenotypes solely based on interpretation of marker expression (tumor panel) or interpretation of marker expression and astir/GMM prediction fractions for known cell type subsets (myeloid and T cell panels) resulting in 13 tumor phenotypes, 5 myeloid phenotypes, 7 CD4^+^ T cell phenotypes, and 9 CD8^+^ T cell phenotypes for downstream analysis.

#### District classification

For district classification, we first detected spatial communities by clustering each cell based on its interactions (Louvain algorithm) defined by the neighborhood graph generated by *steinbock* using the *detectCommunity* function^80^ from *imcRtools*. We set the minimum number of cells per community to 20, 4, and 4 for analysis in the tumor, myeloid, and T cell panels, respectively, and only selected cells of fine-grained phenotypes. Next, we tested different algorithms (hierarchical clustering, agnes, *k*-means) and number of clusters with the *clValid* R package (v.0.7) for clustering each spatial community by its phenotype fractions. This defined 13 (agnes algorithm), 10 (agnes), and 16 (*k*-means) tumor, myeloid, and T cell districts, respectively. This district framework provided us with a measure for the homotypic aggregation of phenotypes in space.

#### Interaction analysis

Pairwise cell-to-cell interaction testing for phenotypes was performed using the *countInteractions* function from the *imcRtools* R/Bioconductor package (v.1.15.2; specifically installed for this analysis). For each image, we computed the average cell type-cell type interaction score (i.e., the fraction of interactions of cell type A that occur with cell type B) using the neighborhood graph generated by *steinbock*. Per panel, we focused on heterotypic interactions of phenotypes (“from”) with broad TME cell types (“to”), thereby complementing the district analysis.

#### Survival analysis

To extract survival-relevant features of expression and phenotypes (non-spatial) as well as districts and interactions (spatial), we used Kaplan-Meier survival estimates from log-rank tests and hazard ratios from multivariable Cox proportional hazard models (Wald test) implemented in the R package *survival* (v.3.5.7). For expression, we extracted mean phenotype-specific min-max normalized expressions per patient across selected markers per panel (tumor panel: CA9, panCK, GPX4, HLADR, NFKB, CyclinD1, HIF1a, p53, cMYC, ICAM1, PDK4, CD73, BRG1, PKM2, PDL1, CD10, IDH2, HIF2a, AuroraKinaseA, Tim3, EGFR, p-S6, VEGFR1, VHL, RB1, p-mTOR, GLUT1, Fumarase, GLUD1/2 + CD44 (from T cell panel data); myeloid panel: MPO, CD16, HLADR, CD68, CD163, CD11c, CD11b, MMP9, CCR2, Arginase, CD33, CD209, CD206, CD15, VISTA, IDO1, PDL1; T cell panel: CD45RA, LAG-3, Tim3, PD1, NKG2A, CXCL13, EOMES, Tbet, FOXP3, GATA3, GITR, CD134, CD278; CD8^+^: CD45RA, LAG-3, Tim3, PD1, NKG2A, CXCL13, EOMES, Tbet, GITR, CD134, CD278, GranB, GranK). We then split these scores by mean and median and used log-rank tests-based estimates to extract relevant features.

For phenotypes and districts, we extracted mean phenotype- or district-specific densities per patient per panel, split these scores by mean and median and used multivariable Cox proportional hazards model-based estimates to extract relevant features. For heterotypic interactions, we extracted mean interaction-specific scores per patient and panel, split these scores by mean and median and used log-rank tests-based estimates to extract relevant features. We excluded the “Unknown” broad TME cell type as interaction partners.

#### PDL1-IMC score

We derived a PDL1-IMC score from PDL1 expression patterns in the myeloid panel. We used GMM (Uncertainty = 0) to detect cells positive or negative for PDL1 and then used a combined positive score of ≥1% of all cells per patient as the threshold for positivity.

#### Disease progression analysis

To extract progression-relevant features of phenotypes (non-spatial) and districts (spatial), we used differential abundance testing implemented in the *edgeR* R/Bioconductor package (v.4.0.2)^81^, following the workflow at https://bioconductor.org/books/release/OSCA/, across clinical (ISUP grade (1/2 versus 3/4), T stage (1/2 versus 3/4), necrosis (absence versus presence)) and immunological (PDL1-IMC score (positive versus negative)) proxies of disease progression. Specifically, we used negative binomial general linear models to model over-dispersed count data, with the count equal to the number of cells per label. After estimating negative binomial and quasi-likelihood dispersions (both with disabled trend), we tested for differential abundance by fitting a quasi-likelihood negative binomial generalized log-linear model and then applying empirical Bayes quasi-likelihood F-tests to the coefficients.

#### Survival prediction machine learning

To perform survival prediction, we preprocessed survival-relevant features following previous reports^31,32^. This included removal of numerical features with more than 30% of missing values and missing value imputation with the minimum value per feature across non-missing samples for interaction features. We standardized all numerical features using Z-score transformation. To reduce collinearity, we removed highly correlated numerical features above 90% Pearson correlation and highly associated categorical features at p<0.001 from chi-square testing. Notably, for clinical features, this led to the selection of age, gender, and T stage and removal of ISUP grade and necrosis status. We then selected patients with complete information across panels including OS data, which resulted in 412 patients for downstream prediction analysis.

We used the statistical machine-learning method *BlockForest*^30^ (v.0.2.6) for survival prediction, which is based on the random survival forest algorithm^33^. This method can consider group structures in multi-modal datasets and has performed well in recent benchmarks^31,32^. We performed 10-fold outer cross-validation and stratified train/test-sets (75-25%) by censoring status (i.e., resulting train and test sets have similar censoring rates). Settings for the *blockfor* function were largely default and included 2000 trees for training, the ‘extratrees’ split rule, 300 sets of random tuning parameter values, 1500 trees for parameter tuning, no block favoring and permutation importance to assess variable importances. Model performance metrics were Integrated Brier scores and Concordance-Index (as proposed by Uno and colleagues^34^). Integrated Brier Scores, also called cumulative prediction error curves, are a measure for predictive calibration and discrimination of survival times and were calculated between the minimum (always 0) and the 0.9 quantile of observed survival times in each test split via the R package *pec* (v.2023.04.12).

#### Spatial CD8^+^ T cell analysis

Based on a previous study^43^, we assigned one of three spatial CD8^+^ patterns to each CD8^+^ T cell in the T cell panel based on the number of tumor cells in direct spatial proximity using the neighborhood graph generated by *steinbock* as follows: *Infiltrating*, ≥ 3 tumor cells; *Bystanding*, 1-2 tumor cells; *Ignored*, 0 tumor cells. Next, we used these spatial CD8^+^ patterns to assign a spatial CD8^+^ group to every patient as follows: *Inflamed* images were identified by a bystanding-to-infiltrating cell ratio of < 7 and ≥ 5 Infiltrating cells. *Excluded* images were defined by a bystanding-to-infiltrating cell ratio of ≥ 7 or < 5 Infiltrating cells. *Cold* images were classified based on a combined count of Infiltrating and Bystanding cells < 6 and < 20 Ignored cells. All patients with the same spatial CD8^+^ group per image were assigned to a respective spatial CD8^+^ group. Patients with images of different spatial CD8^+^ groups had specific classification rules. Patients with both *Inflamed* and *Cold* images were assigned to the *Inflamed* group due to higher Infiltrating and lower Ignored cell counts. Those with *Inflamed* and *Excluded* images were also classified as *Inflamed*, based on stronger association with Infiltrating cell counts. Patients with both *Cold* and *Excluded* samples were assigned to the *Cold* group, reflecting low Bystanding and Ignored cell counts. Patients with images across all spatial CD8^+^ groups were classified as *Excluded*. We compared these spatial groups to number groups, which were based on a tertile split of the average total number of CD8^+^ T cells per patient.

### H&E data

#### Tissue preparation and staining

FFPE tissue sections of 2 µm thickness were deparaffinized in xylene three times for 10 min each wash and rehydrated through a graded alcohol series (ethanol:deionized water, 2 × 100:0 for 5 min; 2 × 96:4, 90:10, 80:20, 70:30 for 3 min each), followed by a 1 min wash with double distilled H₂O. Samples were then stained with hematoxylin (DAKO, Agilent) for 3 min, rinsed in Scots tap water for 1 min, and subsequently dehydrated in 96% ethanol for 1 min. Eosin Y (DAKO, Agilent) was applied for 2 min, followed by another dehydration step in 96% ethanol for 1 min, and three washes in 100% ethanol for 1 min each. Sections were cleared in xylol (2× 1 min) and embedded using Eukitt.

#### Deep learning embedding and classification framework

To predict survival ecotypes for the discovery cohort from 20X magnification H&E-stained images (n = 796 images, 412 patients) consecutive to our IMC data, we used the CLAM toolbox (https://github.com/mahmoodlab/CLAM; Python 3.10.0)^45^, a deep learning-based pipeline for data efficient and weakly supervised pathology tissue analysis, which uses an attention-based multiple instance learning method for classification. After segmentation with custom parameters and patching (patch size = 256), we extracted embeddings with UNI (uni_v1, target patch size = 224, embedding size = 1024, total patch number = 90942)^44^, a self-supervised model for computational pathology. We performed a hyperparameter grid search across different B {4,8,16,32} (number of positive/negative patches to sample for the CLAM model (clam_sb)), learning rates {1e-4, 2e-5, 1e-5, 5e-6} with an adam optimizer, bag losses {ce, svm}, and dropout rates {0.25, 0.3. 0.35} on a single random validation fold by training on 50% of the full training data for a maximum of 30 epochs. Images from the same patient were always assigned to the same train/validation/test splits (80-10-10%). We trained the full model across 10-fold Monte Carlo cross-validation using following hyperparameters: B = 8, learning rate = 1e-5, bag loss = ce, dropout = 0.35 and otherwise default settings.

We used the trained model for survival ecotypes prediction on whole-slide H&E images from the TCGA KIRC database (n = 519 images (512 patients), at 40X magnification), which we download from the Genomic Data Commons portal (downloaded on 14.03.2025). Images were preprocessed as follows: segmentation with provided TCGA parameters and patching (patch size = 512, step size = 512, patch level = 0), and feature extraction with UNI (uni_v1, target patch size = 256)^44^. We selected the survival ecotype with the maximum prediction probability across folds per patient.

#### Embedding clustering

For clustering, patch-level embeddings extracted with UNI (uni_v1, target patch size = 224, embedding size = 1024, total patch number = 90942)^44^ were Z-score normalized and reduced to 274-dimensional embeddings using principial component analysis, which explained 90% of variance. We then tested the *k*-means algorithm on different number of clusters (10-30), evaluated cluster stability using silhouette width, and selected a *k* of 12 for downstream analysis.

### Targeted genomic sequencing

#### Library preparation, quality control and targeted sequencing

Sequencing libraries were prepared using the HaloPlex HS Target Enrichment System (Agilent Technologies) according to the manufacturer’s instructions. The content of the HaloPlex HS gene panel has been described previously^35^. For target enrichment, 100 ng of DNA extracted from FFPE-derived samples was used. The quality of the final libraries was assessed using a Fragment Analyzer (Agilent Technologies). Paired-end sequencing (PE100) was performed on an Illumina NovaSeq 6000 system.

#### Basic data processing

Sequencing of samples from patients of the discovery cohort was performed in two separate efforts. For cohort 1 (n=89 patients)^35^, raw fastq files were downloaded from European Nucleotide Archive (Project: PRJEB26346), all fastq files belonging to the same sample were merged, and subsequently preprocessed according to the description on the European Nucleotide Archive to trim trailing bases particular to the HaloPlexHS protocol and prepare fastq files for read processing based on unique molecular identifiers (UMIs). For cohort 2 (n=95 patients), fastq files were prepared for UMI processing using *umi_tools* (v.1.1.2) and HaloPlexHS protocol-specific bases were trimmed using *fastp* (v.0.23.4). All samples were analyzed with *Sarek* (v.3.4.0)^82^ for UMI consensus read analysis, read trimming and filtering, quality control, and read mapping to the hg38 human reference genome.

#### Mutation calling

Variant calling was performed using Genome Analysis Toolkit (GATK) *Mutect 2* (gatk4-4.2.6.1-1)^83^ using a custom *Snakemake* workflow (https://github.com/ETH-NEXUS/run_gatk_mutect2) and including FFPE-specific filter settings to address the FFPE samples: Mutect2 was run and filtered with setting --f1r2-tar-gz, and FFPE artifacts were flagged using SOBDetector (v.1.0.4). All variants were annotated using *VEP* (build 108)^84^. In cohort 1, multiple tumor samples and a matched normal control were available per patient, and we performed a joint variant call analysis combining the evidence of all samples belonging to the same patient. In cohort 2, matched controls were not available, and we created a custom Panel of Normals based on the 89 normal control samples of cohort 1 that was used as control for the variant calling of cohort 2 tumor samples.

#### CNV calling

For cohort 1, CNV calling and tumor purity estimation were performed using *PureCN* (v.2.10)^85^. Analysis was performed using a custom *Snakemake* workflow (https://github.com/ETH-NEXUS/pureCN_workflow), including *PureCN* analysis and postprocessing as follows: After Sarek preprocessing and as preparation for the CNV calling, single-nucleotide variants were identified using GATK *Mutect 2* in joint mode for all samples belonging to the same patient, with the parameters suggested for PureCN (--genotype-germline-sites true; --genotype-pon-sites true; --interval-padding 75). To exclude potential technical artifacts, variants were filtered, and variants tagged with any of the following terms were excluded: base_qual, contamination, duplicate, fragment, low_allele_frac, n_ratio, map_qual, orientation, position, possible_numt, slippage, strand_bias, strict_strand, weak_evidence. Resulting VCF files were split and processed with *PureCN* scripts. In brief, the amplicon targets were transformed to the hg38 reference genome and processed. For all samples, GC normalized coverage was computed. Normal samples from cohort 1 were used to construct the normal database. The *PureCN* algorithm was then applied to all tumor samples from cohort 1. Of the 178 tumor samples, 167 (94%) passed quality control.

#### Data analysis

For downstream analysis, mutations called by *Mutect 2* across all patients (n=184 patients; 173 with OS information) were further filtered and annotated using following criteria (applied in order): 1) only variants with ≥ 2 reads supporting the variant and a variant allele frequency (VAF) > 5% were retained; 2) only variants with a HIGH or MODERATE impact rating based on ENSEMBL were retained and annotated (splice_acceptor_variant”, “splice_donor_variant”, “stop_gained”, “frameshift_variant”, “stop_lost”, “start_lost”, “inframe_insertion”, “inframe_deletion”, “missense_variant”, “protein_altering_variant”); 3) for cohort 1, mutations found in both tumor samples were considered clonal and otherwise subclonal, whereas due to the lack of paired normal samples in cohort 2, all mutations were considered clonal; 4) high-confidence driver genes, based on data from the Integrative OncoGenomics (IntOGen) pipeline^86^ for ccRCC (n=44; Downloaded on 10.04.2024), were selected. We added focal CNV events for IntOGen driver genes from the *pureCN* pipeline, classified as an AMPLIFICATION or DELETION, to the mutation analysis. Mutational burden per patient was calculated across all genes (after filtering for variants reads and VAF) and included all impact ratings as well as clonal (counted as 1) and subclonal (counted as 0.5) mutations.

Arm-level events were estimated and annotated from CNV events called by *pureCN* across cohort 1 patients (n=82 patients, 79 with OS information) as follows (applied in order): 1) Normalized copy numbers (nCN) were estimated by weighting the copy number values of individual segments based on their base pair lengths relative to the total base pair length of all segments within the same chromosome arm; each arm was then classified into one of three categories (gain (nCN ≥ 3), loss (nCN < 1.5) and otherwise neutral); 2) arm-level events found in both tumor samples were considered clonal and otherwise subclonal; 3) known arm-level events from the TRACERx renal study^17^ and frequent events observed in our study (gains of chr5/7/12p, chr1/2/5/7/8/20q and losses of chr1/3/8/9p, chr4/6/9/13/14/18q) were annotated. CNV burden was estimated across all arm-level events including clonal (counted as 1) and subclonal (counted as 0.5) events.

#### Genomic-like subtypes

Patients were assigned to one of five genomic-like subtypes based on following rules from the TRACERx renal study^17^ (applied in order): 1) presence of >=2 *BAP1*, *PBRM1*, *SETD2*, or *PTEN* mutations (clonal) = “Multiple drivers”; 2) presence of a *BAP1* mutation (clonal/subclonal) was classified as “BAP1 driven”; 3) presence of a *PBRM1* mutation (clonal/subclonal) was classified as “PBRM1 driven” (this subtype encompassed the originally proposed PBRM1>SETD2, PBRM1>PI3K and PBRM1>CNA subtypes, which we did not differentiate due to the lower number of biopsies per patient); 4) presence of only mutant *VHL* (clonal/subclonal) was classified as “VHL monodriver”; 5) absence of *VHL* mutation was classified as “VHL wildtype”.

### MALDI-MSI data

#### Tissue preparation and acquisition

Untargeted MALDI-MSI data for the ccRCC discovery cohort were obtained from a previous study^40^. In brief, tissue preparation was performed as outlined before^87^ and included indium-tin-oxide-coated glass slide mounting and spray-coating of the matrix (10 mg/mL 9-aminoacridine hydrochloride monohydrate matrix (Sigma-Aldrich) in 70% methanol). Acquisition was performed with a 7 T Solarix XR FT-ICR mass spectrometer (Bruker) equipped with a dual ESI-MALDI source and a SmartBeam-II Nd: YAG (355 nm) laser in negative-ion mode. Pixel resolution was 60 µm. MALDI-MSI mass spectra were root mean square normalized with SCiLS (v. 2020b Pro) and a Python 3 pipeline was used for peak picking. After noise filtering, only local maxima were kept as preliminary peaks and subsequently merged and aligned. Peaks that occur in less than 0.5% of the spectra were not considered. For co-registration of the MALDI-MSI measurement regions with H&E images of the same TMA sections, a SPACiAL pipeline was used^88^. Mean peak intensities of tumor regions were extracted for each patient and used for downstream analysis.

#### Data analysis

We used feature annotations from the original study and performed an additional mass search in the KEGG database using the *KEGGREST* R/Bioconductor package (v.1.42.0). After filtering for human and KEGG pathway annotated compounds and removal of m/z values with 0 intensity across all patients, we identified 121 m/z values for downstream analysis. Subsequent data processing included zero-value imputation (replacing zeros with 1/5 of the minimum value), log transformation, batch effect correction via the *mnnCorrect* function from *batchelor* R/Bioconductor package (v.1.18.0), and min-max scaling. We used *SummarizedExperiment* (v.1.32.0) as the data container for downstream analysis (n=443 patients, 370 with OS information).

To identify metabolic programs in our cohort, we employed a two-step clustering and pathway enrichment analysis approach. First, we tested different algorithms (hierarchical clustering, agnes, *k*-means), number of clusters and metrics (including silhouette width and Dunn index) with the *clValid* R package (v.0.7) for grouping the m/z features into metabolic clusters. We selected 17 clusters with *k*-means clustering. Next, we performed pathway enrichment analysis for each cluster using *MetaboAnalyst 6*^89^. For each cluster, we calculated p-values by comparing the min-max expression values for a given cluster to all other clusters with a Student’s *t*-test and used these as input for the “Functional Analysis” module (mass tolerance = 4 ppm, negative ion mode, mummichog algorithm^90^, human KEGG pathway library). We then identified eight distinct metabolic subtypes by clustering the resulting -log10-transformed p-values for each pathway across clusters (“P(Fisher)”, hierarchical clustering) after testing different algorithms, number of clusters, and metrics as described above.

### Bulk transcriptomic sequencing

#### TCGA KIRC data analysis

We downloaded RNA-seq data for the TCGA KIRC cohort (n = 533 patients)^19^ using the *TCGAbiolinks* R/Bioconductor package (v.2.30.0)^91^. We specified the GDC query as follows: Project – TCGA KIRC; Data category – Transcriptome Profiling; Data type – Gene Expression Quantification; Workflow type – STAR counts; Sample type – Primary tumor. We used *SummarizedExperiment* (v.1.32.0) as the data container for downstream analysis. Clinical information, mutation, and survival data were downloaded from the UCSC Xena browser (downloaded on 15.10.2024)^92^. For patients with more than one sample (n=4), we used the median count across samples per patient. For count normalization, the *DESeq2* R/Bioconductor package (v.1.42.0)^93^ was used, which performs size factor-based normalization. Variance-stabilizing transformation was applied to the normalized counts to yield approximately homoscedastic values, which we refer to as expression values. We filtered patients for available survival ecotype prediction information from the H&E deep learning prediction pipeline and performed downstream analysis in 510 patients.

#### Gene signature analysis

All gene signatures used in the current study, except our proposed signatures, were obtained from cited publications. We used all genes for the genomic dedifferentiation signature^47^ (*CD44, COL6A1, DCBLD2, GFPT2, L1CAM, PLAU, PLTP, SEMA3C, TFAP2A, TRNP1*), all genes for the de-clear cell differentiated ccRCC (IM4 subtype) signature^26^, and the top 70 genes for the chromosomal instability signature^46^. We excluded all signature genes that were not detected in the processed datasets.

#### Differential gene expression and gene signature extraction

For differential gene expression, we used the *DESeq2* R/Bioconductor package (v.1.42.0)^93^ package. We filtered for protein-coding genes with at least 10 counts and performed differential expression analysis using the *DESeq* function, which performs size factor-based count normalization, dispersion estimation, and fitting of a negative binomial generalized linear model and computes Wald statistics^93^. Results were filtered based on the mean of normalized counts for each gene and variance-stabilized.

To extract H&E-based gene signatures for the *Poor* and *Favorable* survival ecotypes, we filtered for differentially expressed genes (DEGs) at FDR < 0.05 and log2FoldChange > 1 per comparison and used the intersection of DEGs per group. This resulted in 57 and 47 genes for the *Poor* and *Favorable* groups, respectively **(Table S4)**.

#### Gene set enrichment analysis

Gene set enrichment analysis of Gene Ontology (GO) terms was performed with the *clusterProfiler* R/Bioconductor package (v.4.10.0) per comparison. We specified the minimal and maximal gene set size to be 50 and 1000, respectively, and otherwise used default settings. Per group, we used the intersection of enriched GO terms with an enrichmentScore > 0.

#### Immune cell deconvolution

Immune cell deconvolution of TCGA KIRC RNA-seq data was performed using the quanTIseq^94^ and MCP-counter^95^ methods, implemented in the *immunedeconv* R package (v.2.1.0), by providing a transcripts per million (TPM) normalized gene expression matrix.

#### CPTAC CCRCC data analysis

We downloaded RNA-seq data from the CPTAC cohort from the GDC portal (STAR gene counts, downloaded on 19.05.2025) and metadata from the original publication^23^. We used *SummarizedExperiment* (v.1.32.0) as the data container for downstream analysis. For patients with more than one sample (n=42), we used the median count across samples per patient. We filtered for cases with metadata, resulting in 212 patients. For count normalization, the *DESeq2* R/Bioconductor package (v.1.42.0)^93^ was used as described above.

#### JAVELIN 101 data analysis

We downloaded processed clinical and transcriptomic data from the JAVELIN 101 clinical trial^4^ (n=733 patients) from a previous study^96^. We used *SummarizedExperiment* (v.1.32.0) as the data container for downstream analysis.

#### CheckMate data analysis

We downloaded processed clinical and transcriptomic data from the CheckMate clinical trial (n=311 patients) from the original study^48^. The transcriptomic data contained batch-corrected expression values based on log2-transformed TPM values across three CheckMate cohorts (009, 010, 025). We used *SummarizedExperiment* (v.1.32.0) as the data container for downstream analysis.

#### IMmotion 151 data analysis

We downloaded processed clinical and transcriptomic data from the IMmotion 151 clinical trial^24^ (n=823 patients) from the European Genome-phenome Archive. We used *SummarizedExperiment* (v.1.32.0) as the data container for downstream analysis.

#### Nearest template prediction

To match transcriptomic samples to our survival ecotypes based on our H&E gene signatures, we used the nearest template prediction (NTP) algorithm from the *CMScaller* R package (v.2.0.1)^97,98^. Five independent cohorts (TCGA KIRC, CPTAC, JAVELIN 101, IMmotion 151, and CheckMate) were analyzed. For TCGA KIRC, CPTAC, JAVELIN 101, and IMmotion 151 cohorts, we used TPM values, and, for the CheckMate dataset, we used the provided batch-corrected expression values for log2-transformation (pseudocount of 1) and normalized using the *ematAdjust* function (method = “RLE”). Annotation was performed with the *ntp* function (distance = “cosine”, nPerm = 1000), and patients were assigned to the *Poor* and *Favorable* survival ecotypes based on an FDR threshold (TCGA KIRC, FDR < 0.01; CPTAC, JAVELIN 101, IMmotion 151, FDR < 0.05; CheckMate, FDR < 0.1) and were otherwise called *Medium*.

### Quantification and statistical analyses

Details of used statistical tests are described in the figures and figure legends. If not indicated otherwise, tests were two-tailed. A p value below 0.05 was considered statistically significant. False-discovery rate (FDR) correction for multiple testing was performed using the Benjamini-Hochberg method, and an FDR < 0.1 was considered statistically significant (statistical trends at *P* < 0.05). All other statistics are described in the relevant sections above.

## SUPPLEMENTAL INFORMATION

**Document S1. Supplemental figures S1-S10.**

**Table S1: Antibody panel information for IMC**

*Related to **Methods***

**Table S2: Astir and GMM prediction settings**

*Related to **Methods***

**Table S3: Anova of Cox models for survival ecotypes and basic clinical information**

*Related to **Figure 3 and Figure 5***

**Table S4: Gene signatures for survival ecotypes**

*Related to **Figure 5***

**Table S5: Anova of Cox models for survival ecotypes and IMDC scores**

*Related to **Figure 6***

## RESOURCE AVAILABILITY

### Data and code availability

All data and code will be made publicly available on Zenodo and Github upon publication.

## Data Availability

All data and code will be made publicly available on Zenodo and Github upon publication.

## ACKNOWLEDGMENTS

We thank all patients who donated tumor samples, the B.B. research group for fruitful discussions and feedback, and Genentech, Jack Kuipers, Prof. Axel Karl Walch, and Na Sun for dataset access support. Targeted genomic sequencing was performed at the Genomics Facility Basel, operated by the University of Basel and the Department of Biosystems Science and Engineering (D-BSSE), ETH Zurich.

## AUTHOR CONTRIBUTIONS

*CRediT author statement*

Lasse Meyer: Conceptualization, Methodology, Software, Formal analysis, Investigation, Visualization, Writing - Original Draft, Writing - Review & Editing

Stefanie Engler: Methodology, Investigation

Marlene Lutz: Software, Investigation

Peter Schraml: Data Curation Dorothea

Rutishauser: Data Curation

Anne Bertolini: Software, Investigation

Matthias Lienhard: Software, Investigation

Franziska Singer: Resources

Christian Beisel: Resources

Natalie De Souza: Writing - Original Draft, Writing - Review & Editing

Niko Beerenwinkel: Resources, Supervision

Holger Moch: Conceptualization, Resources, Supervision, Funding acquisition

Bernd Bodenmiller: Conceptualization, Resources, Supervision, Funding acquisition, Writing - Review & Editing

