## Supplemental Information for "A spatial multi-omic portrait of survival outcome for clear cell renal cell carcinoma"

Table of content

1. FIGURES S1-10

2. SUPPLEMENTAL REFERENCES

### 1. FIGURES S1-10

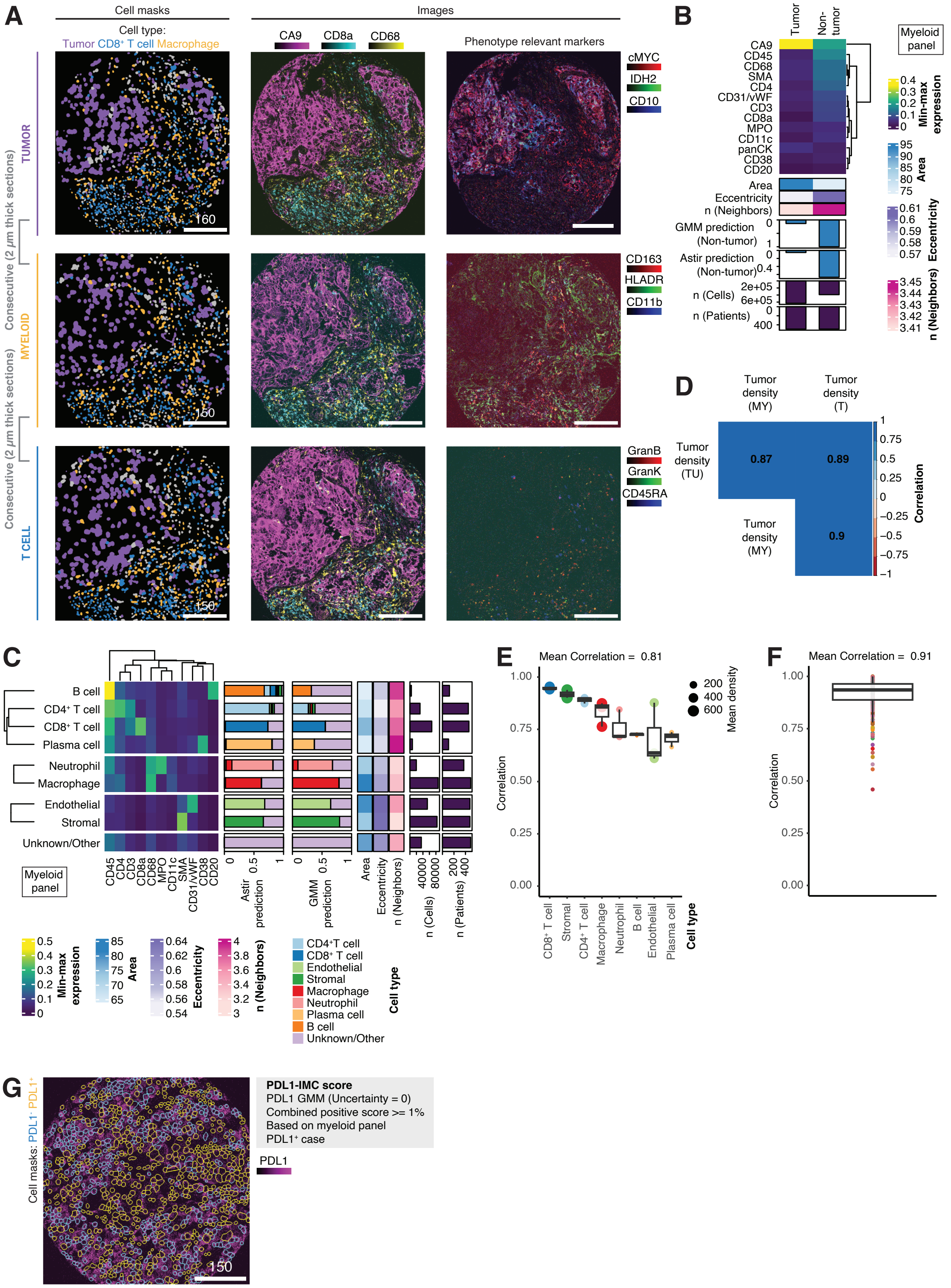

**Figure S1: High-dimensional imaging of the ccRCC ecosystem.** *Related to* ***Figure 1****.*

***(A)*** Representative cell masks of tumor, $CD8^+$ T cells and macrophages (left) and images of celltype- and phenotype-relevant markers (right) from IMC data from consecutive sections across tumor (top), myeloid (center) and T cell (bottom) panels. Scale bars are in µm.

***(B)*** Heatmap of min-max normalized mean marker expression of tumor and non-tumor cells in the myeloid panel as an example. All markers shown were used for graph-based clustering. Spatial single-cell information (area, eccentricity, number of neighbors) averaged per group is indicated. The fractions of non-tumor cells estimated using GMM and *astir*[1] as well as the total number of cells and patients per group are displayed as bar plots.

***(C)*** Heatmap of min-max normalized mean marker expression of broad cell types in the myeloid panel as an example. All markers shown were used for graph-based clustering. Spatial single-cell information (area, eccentricity, number of neighbors), averaged per group is indicated. The fractions of broad cell types estimated using GMM and *astir*[1] as well as the total number of cells and patients per group are displayed as bar plots.

***(D)*** Correlation plot displaying Spearman correlation coefficients for mean tumor cell densities per patient between IMC panels. GMM, Gaussian mixture model.

***(E, F)*** Box plots of Spearman correlation coefficients of mean cell type densities ***(E)*** per cell type and ***(F)*** per patient across IMC panels. Box plots show the median, interquartile range (IQR), and whiskers extend to the most extreme values within 1.5×IQR from the quartiles. Mean correlations are indicated.

***(G)*** Representative image of PDL1 expression with cell masks of $PDL1^+$ and $PDL1^-$ cells based on GMM for a $PDL1^+$ case. Each patient was classified as a $PDL1^+$ patient based on a combined positive score >= 1% of $PDL1^+$ cells (Based on myeloid panel data). Scale bar is 150 µm.

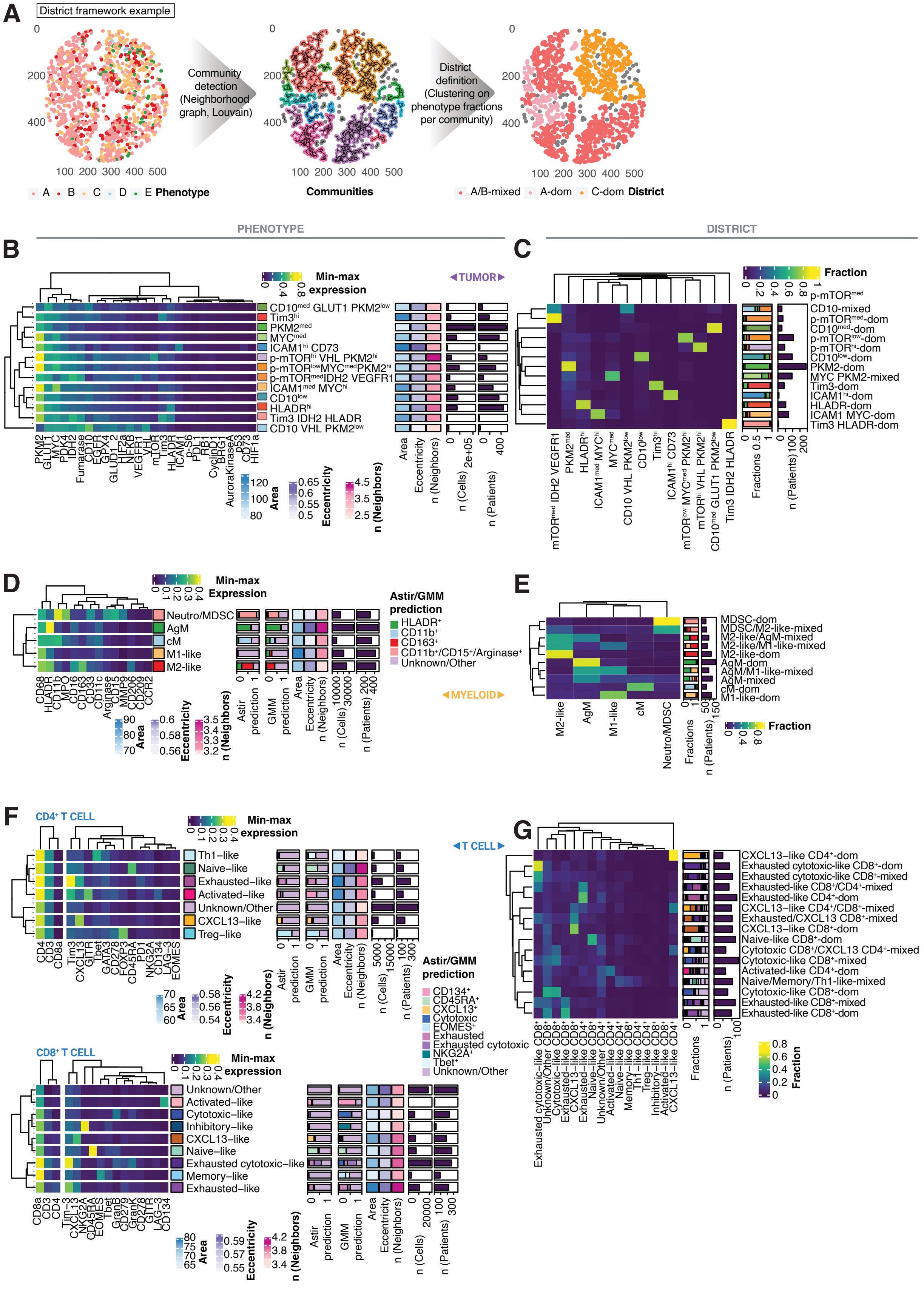
A
District framework example
Community detection (Neighborhood graph, Louvain)
District definition (Clustering on phenotype fractions per community)
Phenotype
Communities
A/B-mixed A-dom C-dom District
PHENOTYPE
DISTRICT
B
C
TUMOR
Min-max expression
Fraction
D
E
MYELOID
Astir/GMM prediction
F
CD4+ T CELL
CD8+ T CELL
G
T CELL

**Figure S2: Phenotypes and districts of tumor, myeloid and T cells.** *Related to* ***Figure 2***.

***(A)*** Schematic showing the workflow for defining districts. Districts provided us with a measure for the homotypic spatial aggregation of phenotypes.

***(B, D, F)*** Heatmap of min-max normalized mean marker expression per phenotype of ***(B)*** tumor, ***(D)*** myeloid and ***(F)*** $CD4^+$ (top) and $CD8^+$ (bottom) T cells. Data for each cellular compartment were from the respective antibody panel. All markers shown were used for graph-based clustering in each cellular compartment. Spatial single-cell information (area, eccentricity, number of neighbors), averaged per cluster is indicated. The fractions of phenotypes estimated using GMM and *astir*[1] as well as the total number of cells and patients per group are displayed as bar plots.

***(C, E, G)*** Heatmap of phenotype fractions per district of ***(C)*** tumor, ***(E)*** myeloid and ***(G)*** T cells. Districts were defined separately for each cellular compartment. The fractions of phenotypes and the total number of patients per group are displayed as bar plots.

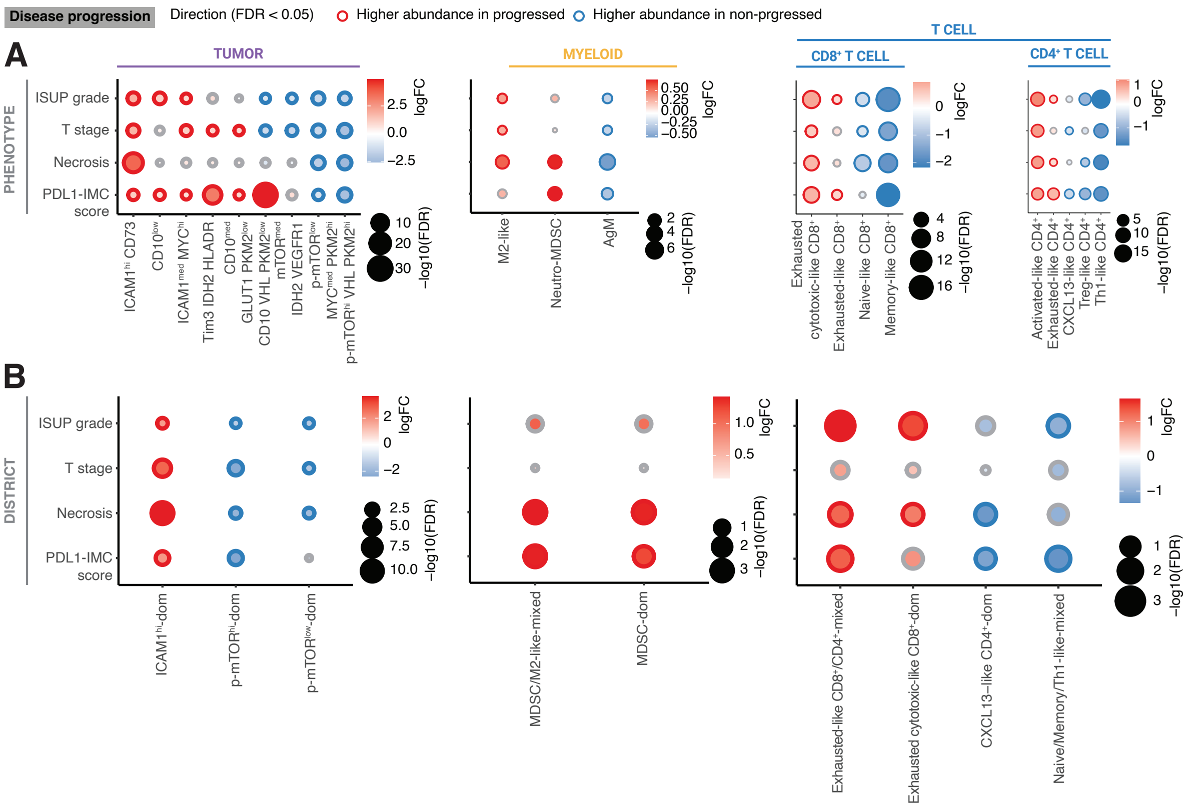


**Figure S3: Progression-associated ecosystem features in ccRCC.** *Related to* ***Figure 2****.*

***(A, B)*** Bubble plots of differentially abundant ***(A)*** phenotypes and ***(B)*** districts of tumor cells (left), myeloid cells (center), and T cells (right) for ISUP grade, T stage, necrosis, and PDL1-IMC score. Bubble color, outline, and size reflect log-fold changes, abundance direction, and significance, respectively. Only features differentially abundant in at least two progression categories are shown (FDR < 0.05).

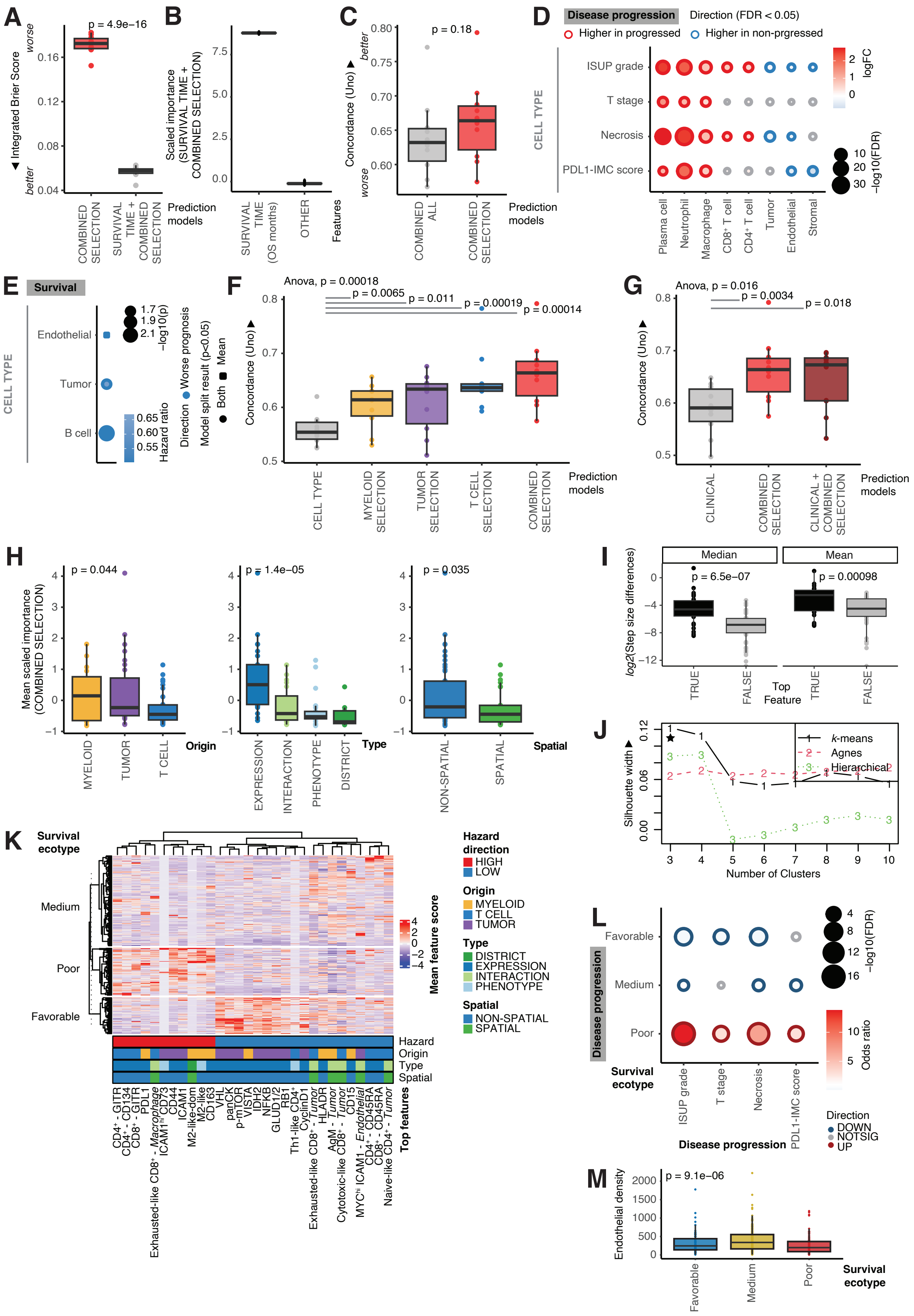
A
B
C
D
Disease progression
Direction (FDR < 0.05)
Higher in progressed
Higher in non-prgressed
E
Survival
F
G
H
I
J
K
L
M
Integrated Brier Score
Scaled importance (SURVIVAL TIME + COMBINED SELECTION)
Concordance (Uno)
Prediction models
Features
CELL TYPE
Mean scaled importance (COMBINED SELECTION)
log2(Step size differences)
Silhouette width
Number of Clusters
Survival ecotype
Mean feature score
Hazard direction
Origin
Type
Spatial
Top features
Endothelial density

**Figure S4: Survival prediction framework underlines ecosystem feature importance.** *Related to* ***Figure 3****.*

***(A)*** Box plots of Integrated Brier Scores across 10-fold outer cross-validation comparing models with selected survival-relevant ecosystem features across compartments (COMBINED SELECTION) and the same models including the survival time as a feature (SURVIVAL TIME + COMBINED SELECTION) (n=412 patients). Lower Integrated Brier Scores indicate better calibration and discrimination of predicted survival times. Individual points show the test estimate for a single cross-validation run. Box plots show the median, IQR, and whiskers extend to the most extreme values within 1.5×IQR from the quartiles. The *P* values were determined using unpaired one-sided t-tests versus relevant baselines.

***(B)*** Box plots of scaled importances of the indicated feature classes in 10-fold outer cross-validation in the models with selected survival-relevant ecosystem features across compartments and survival time (SURVIVAL TIME + COMBINED SELECTION).

***(C, F, G)*** Box plots of Concordance (Uno) across 10-fold outer cross-validation comparing ***(C)*** a model with all ecosystem features (COMBINED ALL) versus a model with selected survival-relevant ecosystem features across compartments (COMBINED SELECTION), ***(F)*** a model with broad cell type densities only (CELL TYPE) versus models with selected survival-relevant ecosystem features within and across compartments (TUMOR SELECTION, T CELL SELECTION, MYELOID SELECTION, COMBINED SELECTION) and ***(G)*** a model with basic clinical features (age, gender, T stage) versus a model with selected survival-relevant ecosystem features (COMBINED SELECTION) and a model combining basic clinical features and ecosystem features (CLINICAL + COMBINED SELECTION) (n=412 patients). Higher Concordance scores indicate better patient risk ranking based on the predicted survival times. The *P* values were determined using unpaired one-sided t-tests versus relevant baselines and one-way ANOVA across groups.

***(D)*** Bubble plots of differentially abundant broad cell types for ISUP grade, T stage, necrosis, and PDL1-IMC score. Bubble color, outline and size reflect log-fold changes, abundance direction and significance, respectively. Only cell types differentially abundant in at least two progression categories are shown (FDR < 0.05).

***(E)*** Bubble plots of survival-relevant broad cell types. Bubble color indicates cox model hazard ratios. Bubble outline, shape and size reflect prognostic direction, model results for the indicated model splits and significance, respectively. Only features with at least one significant association are shown (*P* < 0.05).

***(H)*** Box plots of mean scaled importance of the indicated classes of features for 10-fold outer cross-validation in models with selected survival-relevant ecosystem features across compartments (COMBINED SELECTION). Features are classified according to origin (left), feature type (centre), and whether they are spatial/non-spatial (right). The *P* values were determined using one-way ANOVA across groups.

***(I)*** Box plots of median (left) and mean (right) step-size differences (log2-transformed) between top features and other features. The *P* values were determined using one-way ANOVA across groups.

***(J)*** Silhouette width for different numbers of clusters (n=3-10) and the indicated algorithms. *K*-means clustering and three clusters were selected for downstream analysis and are marked by a star.

***(K)*** Heatmap of mean top feature scores per patient for survival ecotypes. Prognostic directions, feature origin, feature type and spatial annotations are shown.

***(L)*** Bubble plots of association of survival ecotypes with ISUP grade, T stage, necrosis, and PDL1-IMC score. Bubble color, outline and size reflect odds ratio from Fisher exact testing, association direction (FDR < 0.1) and significance, respectively.

***(M)*** Box plots of mean endothelial cell density (in cells/$mm^2$) per patient across survival ecotypes. *P* values were determined using Kruskal-Wallis tests.

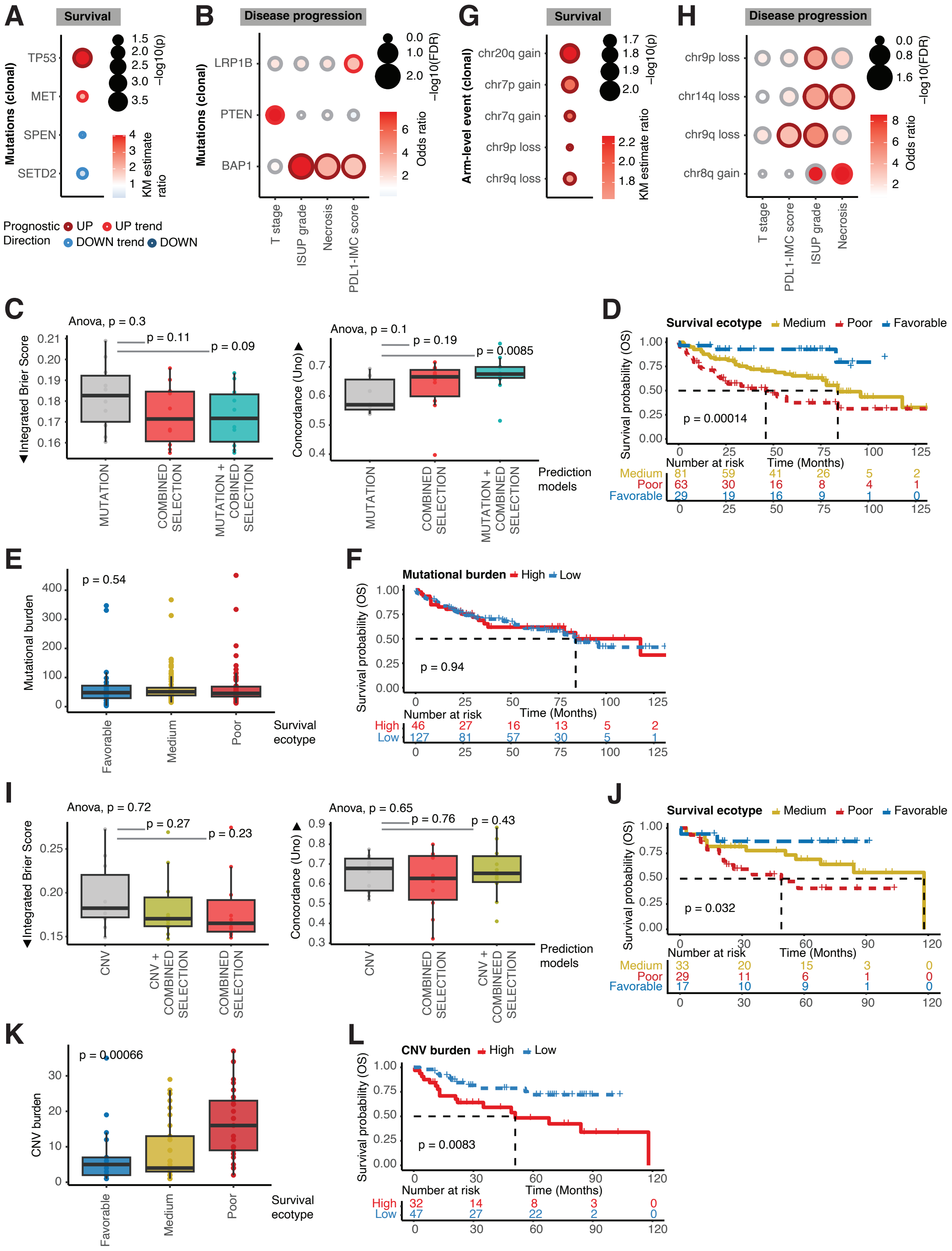
A
Survival
Mutations (clonal)
TP53
MET
SPEN
SETD2
−log10(p)
KM estimate ratio
Prognostic Direction
UP
UP trend
DOWN trend
DOWN
B
Disease progression
LRP1B
PTEN
BAP1
T stage
ISUP grade
Necrosis
PDL1-IMC score
−log10(FDR)
Odds ratio
G
Survival
Arm-level event (clonal)
chr20q gain
chr7p gain
chr7q gain
chr9p loss
chr9q loss
H
Disease progression
chr9p loss
chr14q loss
chr9q loss
chr8q gain
C
Anova, p = 0.3
p = 0.11
p = 0.09
Integrated Brier Score
MUTATION
COMBINED SELECTION
MUTATION + COBINED SELECTION
Anova, p = 0.1
p = 0.19
p = 0.0085
Concordance (Uno)
MUTATION
COMBINED SELECTION
MUTATION + COMBINED SELECTION
Prediction models
D
Survival ecotype
Medium
Poor
Favorable
Survival probability (OS)
p = 0.00014
Number at risk
Time (Months)
E
p = 0.54
Mutational burden
Favorable
Medium
Poor
Survival ecotype
F
Mutational burden
High
Low
p = 0.94
Number at risk
Time (Months)
I
Anova, p = 0.72
p = 0.27
p = 0.23
Integrated Brier Score
CNV
CNV + COMBINED SELECTION
COMBINED SELECTION
Anova, p = 0.65
p = 0.76
p = 0.43
Concordance (Uno)
CNV
COMBINED SELECTION
CNV + COMBINED SELECTION
Prediction models
J
Survival ecotype
Medium
Poor
Favorable
p = 0.032
Number at risk
Time (Months)
K
p = 0.00066
CNV burden
Favorable
Medium
Poor
Survival ecotype
L
CNV burden
High
Low
p = 0.0083
Number at risk
Time (Months)

**Figure S5: Genomic landscape of ccRCC.** *Related to* ***Figure 4****.*

***(A, B, G, H)*** Bubble plots of ***(A, B)*** mutations (clonal) and ***(G, H)*** CNV arm-level events (clonal) associated with ***(A, G)*** overall survival (OS) and ***(B, H)*** with ISUP grade, T stage, necrosis, and PDL1-IMC score. Bubble color, outline and size reflect Kaplan-Meier estimate/odds ratios, association direction and significance, respectively. Only mutations and arm-level events with at least one significant OS and two disease progression category associations are shown (FDR < 0.1; trends at $P < 0.05$).

***(C, I)*** Box plots of Integrated Brier Scores (left) and Concordance (Uno) (right) across 10-fold outer cross-validation comparing ***(C)*** a model with survival-relevant mutation features (MUTATION) versus a model with ecosystem features (COMBINED SELECTION) and a model with both feature types (MUTATION + COMBINED SELECTION) (n=173 patients) and ***(I)*** a model with survival-relevant CNV arm-level features (CNV) versus a model with ecosystem features (COMBINED SELECTION) and a model with both feature types (CNV + COMBINED SELECTION) (n=79 patients). Lower Integrated Brier Scores indicate better predictive calibration and discrimination. Higher Concordance scores indicate better patient risk ranking based on the predicted survival times. Individual points show the test estimate for a single cross-validation run. Box plots show the median, IQR, and whiskers extend to the most extreme values within 1.5×IQR from the quartiles. The *P* values were determined using unpaired one-sided t-tests versus relevant baselines and one-way ANOVA across groups.

***(D, F)*** Kaplan-Meier curves of OS for ***(D)*** survival ecotypes and ***(F)*** mutational burden in patients with mutation information (n=173 patients). *P* values were estimated using a log-rank test.

***(E, K)*** Box plots of ***(E)*** mutational burden (n=173 patients) and ***(K)*** CNV arm-level burden (n=74 patients) across survival ecotypes. *P* values were determined using Kruskal-Wallis tests. CNV, copy-number variation

***(J, L)*** Kaplan-Meier curves of OS for ***(J)*** survival ecotypes and ***(L)*** CNV burden in patients with CNV information (n=79 patients). *P* values were estimated using a log-rank test. CNV, copy-number variation

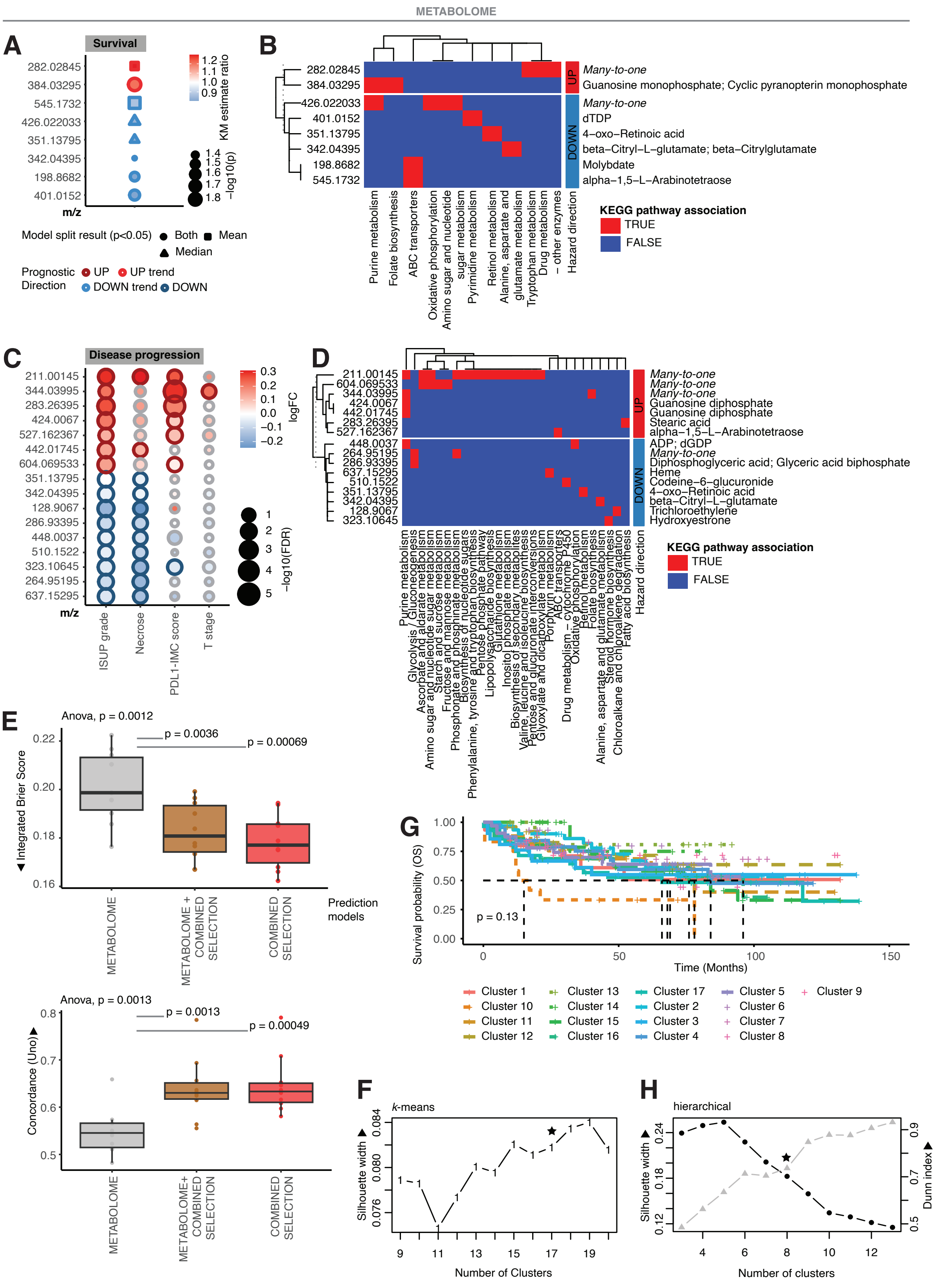
METABOLOME
A
Survival
282.02845
384.03295
545.1732
426.022033
351.13795
342.04395
198.8682
401.0152
m/z
KM estimate ratio
1.2
1.1
1.0
0.9
−log10(p)
1.4
1.5
1.6
1.7
1.8
Model split result (p<0.05) Both Mean Median
Prognostic Direction UP UP trend DOWN trend DOWN
B
UP
DOWN
Many-to-one
Guanosine monophosphate; Cyclic pyranopterin monophosphate
Many-to-one
dTDP
4-oxo-Retinoic acid
beta-Citryl-L-glutamate; beta-Citrylglutamate
Molybdate
alpha-1,5-L-Arabinotetraose
Purine metabolism
Folate biosynthesis
ABC transporters
Oxidative phosphorylation
Amino sugar and nucleotide sugar metabolism
Pyrimidine metabolism
Retinol metabolism
Alanine, aspartate and glutamate metabolism
Tryptophan metabolism
Drug metabolism – other enzymes
Hazard direction
KEGG pathway association
TRUE
FALSE
C
Disease progression
211.00145
344.03995
283.26395
424.0067
527.162367
442.01745
604.069533
351.13795
342.04395
128.9067
286.93395
448.0037
510.1522
323.10645
264.95195
637.15295
m/z
ISUP grade
Necrose
PDL1-IMC score
T stage
logFC
0.3
0.2
0.1
0.0
−0.1
−0.2
−log10(FDR)
1
2
3
4
5
D
211.00145
604.069533
344.03995
424.0067
442.01745
283.26395
527.162367
448.0037
264.95195
286.93395
637.15295
510.1522
351.13795
342.04395
128.9067
323.10645
UP
DOWN
Many-to-one
Many-to-one
Many-to-one
Guanosine diphosphate
Guanosine diphosphate
Stearic acid
alpha-1,5-L-Arabinotetraose
ADP; dGDP
Many-to-one
Diphosphoglyceric acid; Glyceric acid biphosphate
Heme
Codeine-6-glucuronide
4-oxo-Retinoic acid
beta-Citryl-L-glutamate
Trichloroethylene
Hydroxyestrone
Purine metabolism
Glycolysis / Gluconeogenesis
Ascorbate and aldarate metabolism
Amino sugar and nucleotide sugar metabolism
Starch and sucrose metabolism
Fructose and mannose metabolism
Phosphonate and phosphinate metabolism
Biosynthesis of nucleotide sugars
Phenylalanine, tyrosine and tryptophan biosynthesis
Pentose phosphate pathway
Lipopolysaccharide biosynthesis
Glutathione metabolism
Inositol phosphate metabolism
Biosynthesis of secondary metabolites
Valine, leucine and isoleucine biosynthesis
Pentose and glucuronate interconversions
Glyoxylate and dicarboxylate metabolism
Porphyrin metabolism
ABC transporters
Drug metabolism – cytochrome P450
Oxidative phosphorylation
Retinol metabolism
Folate biosynthesis
Alanine, aspartate and glutamate metabolism
Steroid hormone biosynthesis
Chloroalkane and chloroalkene degradation
Fatty acid biosynthesis
Hazard direction
KEGG pathway association
TRUE
FALSE
E
Anova, p = 0.0012
p = 0.0036
p = 0.00069
Integrated Brier Score
0.22
0.20
0.18
0.16
METABOLOME
METABOLOME + COMBINED SELECTION
COMBINED SELECTION
Prediction models
Anova, p = 0.0013
p = 0.0013
p = 0.00049
Concordance (Uno)
0.8
0.7
0.6
0.5
METABOLOME
METABOLOME+ COMBINED SELECTION
COMBINED SELECTION
G
Survival probability (OS)
1.00
0.75
0.50
0.25
0.00
p = 0.13
0
50
100
150
Time (Months)
Cluster 1
Cluster 10
Cluster 11
Cluster 12
Cluster 13
Cluster 14
Cluster 15
Cluster 16
Cluster 17
Cluster 2
Cluster 3
Cluster 4
Cluster 5
Cluster 6
Cluster 7
Cluster 8
Cluster 9
F
k-means
Silhouette width
0.084
0.080
0.076
9
11
13
15
17
19
Number of Clusters
H
hierarchical
Silhouette width
0.24
0.18
0.12
Dunn index
0.9
0.7
0.5
4
6
8
10
12
Number of clusters

**Figure S6: Metabolic landscape of ccRCC.** *Related to* ***Figure 4****.*

***(A)*** Bubble plots of analyte species with the indicated m/z values associated with OS. Bubble color indicates Kaplan-Meier estimate ratios. Bubble outline, shape and size reflect prognostic direction, model split results and significance, respectively. Only m/z values with at least one significant association are shown (FDR < 0.1; trends at p < 0.05).

***(B, D)*** Heatmap of KEGG pathway associations of ***(B)*** survival-relevant and ***(D)*** progression-relevant species with the indicated m/z values, split by prognostic direction. Potential metabolite matches for each m/z value are shown.

***(C)*** Bubble plots of analyte species with the indicated m/z values association with ISUP grade, T stage, necrosis, and PDL1-IMC score. Bubble color, outline and size reflect log-fold chances from Wilcoxon testing, association direction and significance, respectively. Only m/z values with at least two significant associations are shown (FDR < 0.05).

***(E)*** Box plots of Integrated Brier Scores (top) and Concordance (Uno) (bottom) across 10-fold outer cross-validation comparing a model with survival-relevant metabolome features (METABOLOME) versus a model with ecosystem features (COMBINED SELECTION) and a model with both feature types (METABOLOME + COMBINED SELECTION) (n=375 patients). Lower Integrated Brier Scores indicate better predictive calibration and discrimination. Concordance scores indicate better patient risk ranking based on the predicted survival times. Individual points show the test estimate for a single cross-validation run. Box plots show the median, IQR, and whiskers extend to the most extreme values within 1.5×IQR from the quartiles. The *P* values were determined using unpaired one-sided t-tests versus relevant baselines and one-way ANOVA across groups.

***(F)*** Silhouette width for different numbers of clusters (n=9-20) for the *K*-means algorithm. 17 clusters were selected for downstream analysis and are marked by a star.

***(G)*** Kaplan-Meier curves of OS for metabolic clusters (n=382 patients). *P* values were estimated using a log-rank test.

***(H)*** Silhouette width (left axis; black) and Dunn index (right axis; grey) for different numbers of clusters (n=3-13) for the hierarchical clustering algorithm. 8 clusters were selected for downstream analysis and are marked by a star.

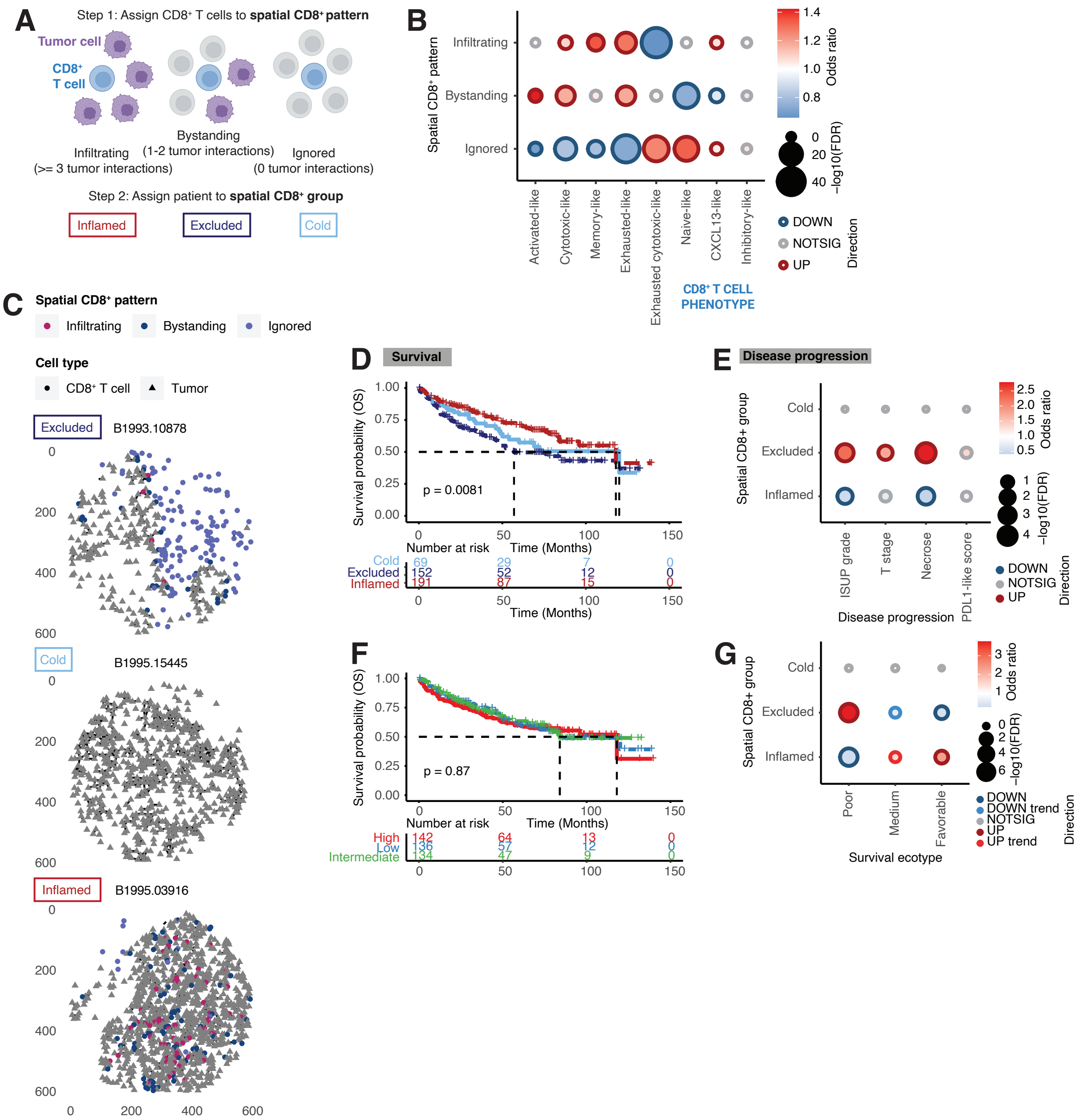


**Figure S7: The spatial location of CD8⁺ T cells influences prognosis and links to ecotypes in ccRCC.** *Related to **Figure 4**.*

***(A)*** Schematic illustrating the assignments of spatial CD8⁺ patterns to CD8⁺ T cells based on interactions with tumor cells, and the assignment of patients to spatial CD8⁺ groups based on the ratio between Bystanding/Infiltrating cells and the total number of Infiltrating cells (Adapted from Meyer et al.[2]).

***(B)*** Bubble plots of association of spatial CD8⁺ patterns with CD8⁺ T cell phenotypes.

Bubble color, outline and size reflect odds ratio from Fisher exact testing, association direction and significance (FDR < 0.1), respectively.

***(C)*** Representative spatial plot of cell centroids for each spatial $CD8^+$ group. Cell color and shape indicate spatial patterns and cell types, respectively.

***(D, F)*** Kaplan-Meier curves of OS for ***(D)*** spatial $CD8^+$ groups and ***(F)*** number groups (based on a tertile split of the total numbers of $CD8^+$ T cells per patient) (n=412 patients). *P* values were estimated using a log-rank test.

***(E, G)*** Bubble plots of association of spatial $CD8^+$ groups with ***(E)*** ISUP grade, T stage, necrosis, PDL1-IMC score and ***(F)*** survival ecotypes. Bubble color, outline and size reflect odds ratio from Fisher exact testing, association direction and significance (FDR < 0.1; trends at $P < 0.05$), respectively.

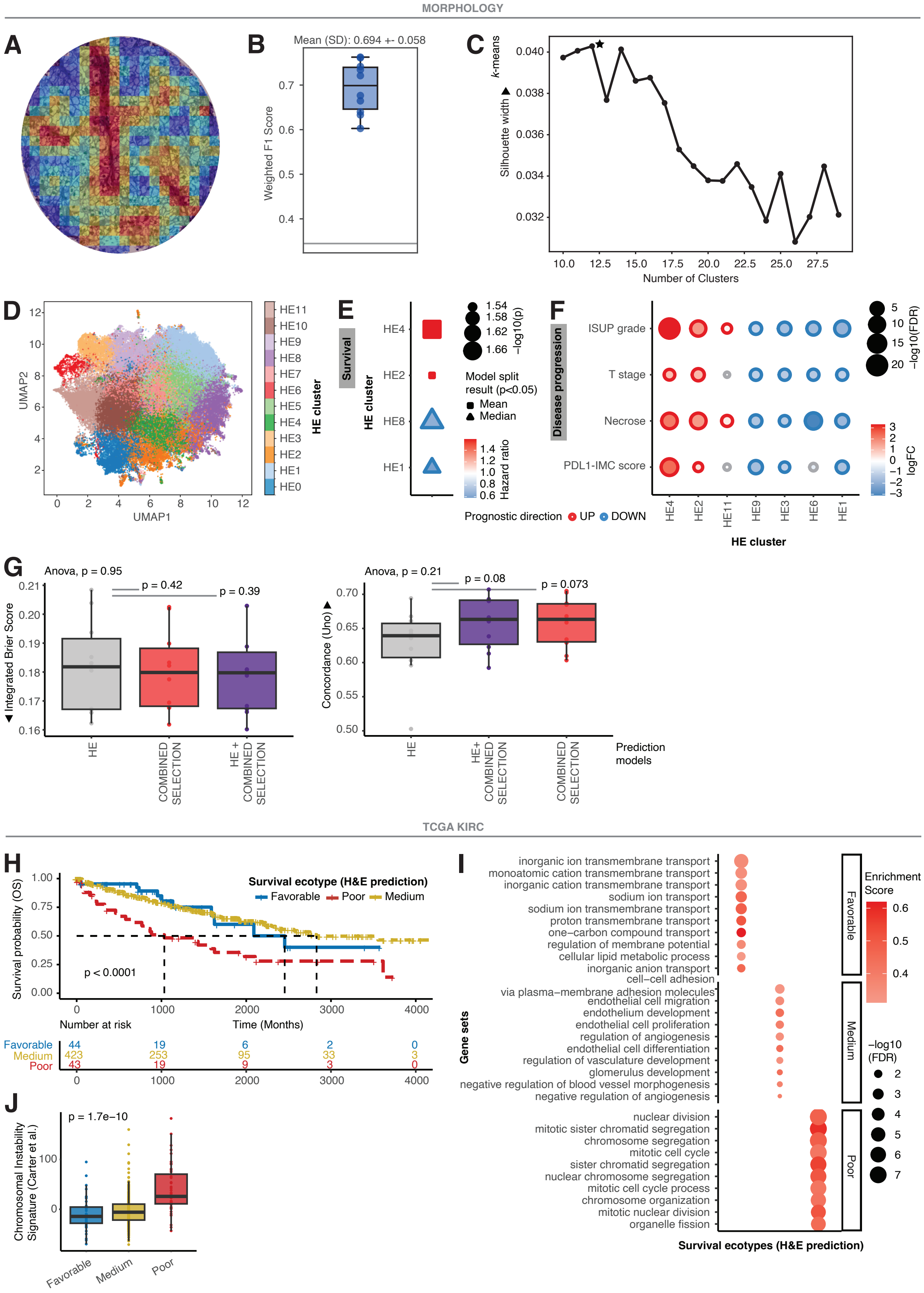
MORPHOLOGY
A
B
Mean (SD): 0.694 +- 0.058
Weighted F1 Score
C
k-means
▲ Silhouette width
Number of Clusters
D
UMAP1
UMAP2
HE cluster
HE11
HE10
HE9
HE8
HE7
HE6
HE5
HE4
HE3
HE2
HE1
HE0
E
Survival
HE cluster
HE4
HE2
HE8
HE1
−log10(p)
Model split result (p<0.05)
Mean
Median
Hazard ratio
Prognostic direction UP DOWN
F
Disease progression
ISUP grade
T stage
Necrose
PDL1-IMC score
−log10(FDR)
logFC
HE cluster
G
Anova, p = 0.95
p = 0.42
p = 0.39
▼ Integrated Brier Score
HE
COMBINED SELECTION
HE + COMBINED SELECTION
Anova, p = 0.21
p = 0.08
p = 0.073
Concordance (Uno) ▲
HE
HE+ COMBINED SELECTION
COMBINED SELECTION
Prediction models
TCGA KIRC
H
Survival ecotype (H&E prediction)
Favorable
Poor
Medium
Survival probability (OS)
p < 0.0001
Time (Months)
Number at risk
Favorable 44 19 6 2 0
Medium 423 253 95 33 3
Poor 43 19 9 3 0
I
inorganic ion transmembrane transport
monoatomic cation transmembrane transport
inorganic cation transmembrane transport
sodium ion transport
sodium ion transmembrane transport
proton transmembrane transport
one−carbon compound transport
regulation of membrane potential
cellular lipid metabolic process
inorganic anion transport
cell−cell adhesion
via plasma−membrane adhesion molecules
endothelial cell migration
endothelium development
endothelial cell proliferation
regulation of angiogenesis
endothelial cell differentiation
regulation of vasculature development
glomerulus development
negative regulation of blood vessel morphogenesis
negative regulation of angiogenesis
nuclear division
mitotic sister chromatid segregation
chromosome segregation
mitotic cell cycle
sister chromatid segregation
nuclear chromosome segregation
mitotic cell cycle process
chromosome organization
mitotic nuclear division
organelle fission
Gene sets
Favorable
Medium
Poor
Enrichment Score
−log10 (FDR)
Survival ecotypes (H&E prediction)
J
p = 1.7e−10
Chromosomal Instability Signature (Carter et al.)
Favorable
Medium
Poor

**Figure S8: Standard pathology images contain ccRCC survival ecotype information.** *Related to* ***Figure 5****.*

***(A)*** Representative H&E image with attention scores (red = high attention, blue = low attention) from deep learning-based classification workflow.

***(B)*** Box plot of weighted F1 test scores across 10-fold Monte Carlo cross-validation for all survival ecotypes. Mean and SD are indicated. Box plots show the median, IQR, and whiskers extend to the most extreme values within 1.5×IQR from the quartiles.

***(C)*** Silhouette width scores for different numbers of clusters (n=10-30) for the *K*-means algorithm. 12 clusters were selected for downstream analysis and are marked by a star.

***(D)*** Two-dimensional UMAP representation of UNI[3]-extracted HE patch embeddings highlighted by cluster. Each dot represents one patch.

***(E)*** Bubble plots of HE clusters associated with OS. Bubble color indicates Cox model hazard ratios. Bubble outline, shape and size reflect prognostic direction, model split results and significance, respectively. Only HE clusters with at least one significant association are shown ($P < 0.05$).

***(F)*** Bubble plots of differentially abundant HE clusters for ISUP grade, T stage, necrosis, and PDL1-IMC score. Bubble color, outline, and size reflect log-fold changes, abundance direction, and significance, respectively. Only features differentially abundant in at least two progression categories are shown (FDR < 0.05).

***(G)*** Box plots of Integrated Brier Scores (left) and Concordance (Uno) (right) across 10-fold outer cross-validation comparing a model with survival-relevant HE features (HE) versus a model with ecosystem features (COMBINED SELECTION) and a model with both feature types (HE + COMBINED SELECTION) (n=412 patients). Lower Integrated Brier Scores indicate better predictive calibration and discrimination. Concordance scores indicate better patient risk ranking based on the predicted survival times. Individual points show the test estimate for a single cross-validation run. Box plots show the median, IQR, and whiskers extend to the most extreme values within 1.5×IQR from the quartiles. The *P* values were determined using unpaired one-sided t-tests versus relevant baselines and one-way ANOVA across groups.

***(H)*** Kaplan-Meier curves of OS for H&E-predicted survival ecotypes in the TCGA KIRC[4] cohort (n=510 patients with H&E data and OS information). *P* values were estimated using a log-rank test.

***(I)*** Bubble plots of the top 10 enriched gene sets (based on FDR) per H&E-predicted survival ecotype in the TCGA KIRC cohort. Bubble color and size reflect enrichment scores and significance, respectively. Only overlapping gene sets i.e., those significantly enriched in all comparisons for a given ecotype, are shown.

***(J)*** Box plots of chromosomal instability signature[5] scores between H&E-predicted survival ecotypes in the TCGA KIRC cohort. *P* values were determined using Kruskal-Wallis tests.

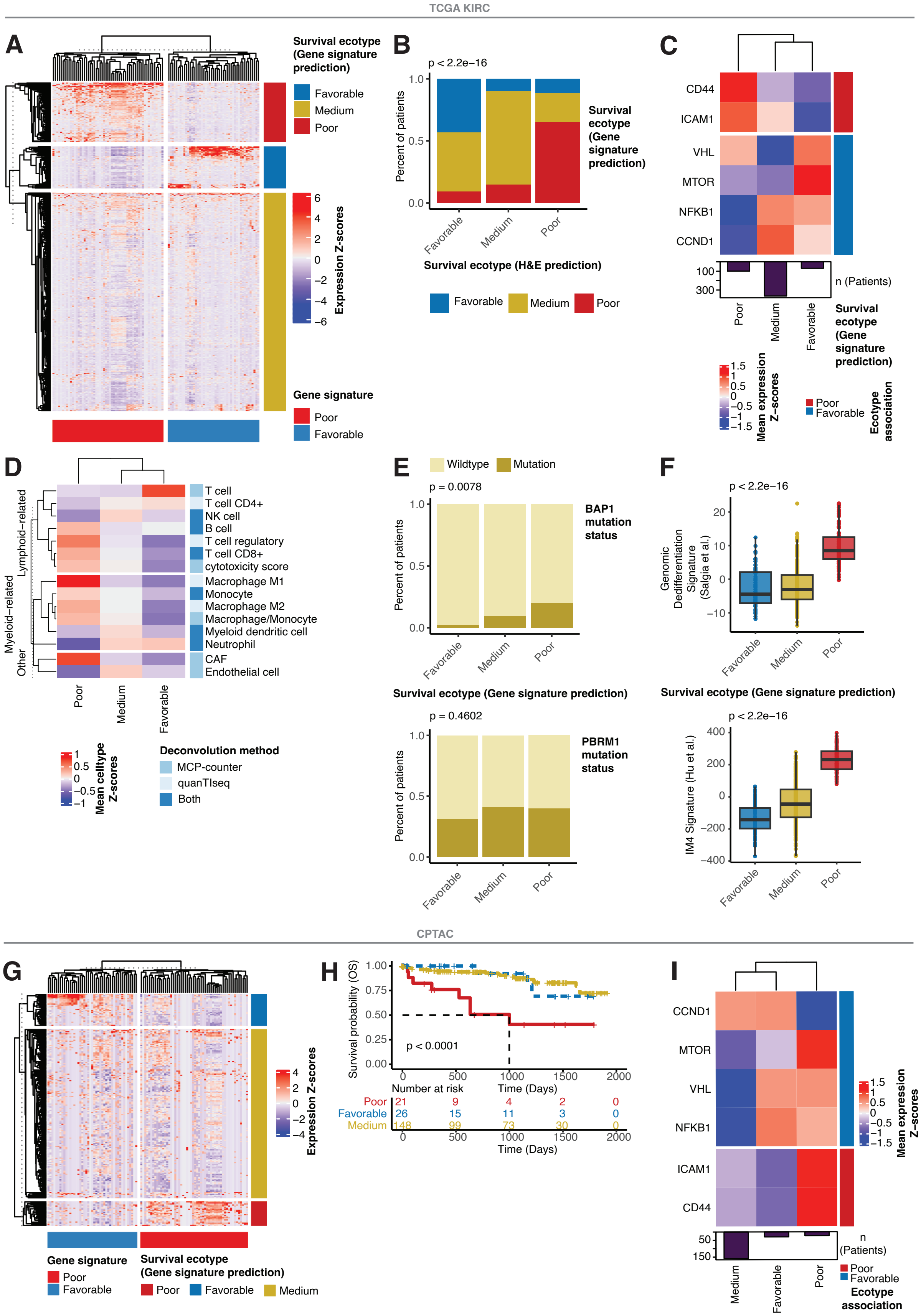

TCGA KIRC
A
Survival ecotype (Gene signature prediction)
Favorable
Medium
Poor
6
4
2
0
−2
−4
−6
Expression Z-scores
Gene signature
Poor
Favorable
B
p < 2.2e−16
1.0
0.5
0.0
Percent of patients
Favorable
Medium
Poor
Survival ecotype (Gene signature prediction)
Survival ecotype (H&E prediction)
Favorable
Medium
Poor
C
CD44
ICAM1
VHL
MTOR
NFKB1
CCND1
100
300
n (Patients)
Poor
Medium
Favorable
Survival ecotype (Gene signature prediction)
1.5
1
0.5
0
−0.5
−1
−1.5
Mean expression Z-scores
Poor
Favorable
Ecotype association
D
Lymphoid−related
Myeloid−related
Other
T cell
T cell CD4+
NK cell
B cell
T cell regulatory
T cell CD8+
cytotoxicity score
Macrophage M1
Monocyte
Macrophage M2
Macrophage/Monocyte
Myeloid dendritic cell
Neutrophil
CAF
Endothelial cell
Poor
Medium
Favorable
1
0.5
0
−0.5
−1
Mean celltype Z-scores
Deconvolution method
MCP-counter
quanTIseq
Both
E
Wildtype
Mutation
p = 0.0078
1.0
0.5
0.0
Percent of patients
BAP1 mutation status
Favorable
Medium
Poor
Survival ecotype (Gene signature prediction)
p = 0.4602
PBRM1 mutation status
F
p < 2.2e−16
20
10
0
−10
Genomic Dedifferentiation Signature (Salgia et al.)
Favorable
Medium
Poor
Survival ecotype (Gene signature prediction)
p < 2.2e−16
400
200
0
−200
−400
IM4 Signature (Hu et al.)
CPTAC
G
4
2
0
−2
−4
Expression Z-scores
Gene signature
Poor
Favorable
Survival ecotype (Gene signature prediction)
Poor
Favorable
Medium
H
1.00
0.75
0.50
0.25
0.00
Survival probability (OS)
p < 0.0001
0
500
1000
1500
2000
Time (Days)
Number at risk
Poor 21 9 4 2 0
Favorable 26 15 11 3 0
Medium 148 99 73 30 0
I
CCND1
MTOR
VHL
NFKB1
ICAM1
CD44
50
150
n (Patients)
Medium
Favorable
Poor
1.5
1
0.5
0
−0.5
−1
−1.5
Mean expression Z-scores
Poor
Favorable
Ecotype association

**Figure S9: H&E based gene signature predicts survival ecotypes.** *Related to* ***Figure 5****.*

***(A)*** Heatmap of expression Z-scores of gene signatures of the poor and favorable survival ecotypes derived from H&E prediction and split by NTP-based gene-signature-predicted survival ecotype results in the TCGA KIRC cohort (n=533 patients with H&E data and OS information). Patients were assigned to the *Poor* and *Favorable* survival ecotypes based on an FDR threshold (FDR < 0.01) and otherwise called *Medium outcome*.

***(B)*** Stacked bar plots of gene-signature predicted survival ecotypes between H&E-predicted survival ecotypes in the TCGA KIRC cohort. *P* values were determined by chi-square testing.

***(C)*** Heatmap of mean expression Z-scores for selected genes across gene-signature-predicted survival ecotypes in the TCGA KIRC cohort. Number of patients per group are shown, and genes are annotated by their protein-level ecotype association in the discovery cohort.

***(D)*** Heatmap of mean Z-scores scores of the indicated cell types across gene-signature-predicted survival ecotypes in the TCGA KIRC cohort. Cell type scores were determined by deconvolution of transcriptomic data using two different methods[6,7]. Rows are split by broad cell lineages.

***(E)*** Stacked bar plots of BAP1 (top) and PBRM1 (bottom) mutation status between gene-signature-predicted survival ecotypes in the TCGA KIRC cohort. *P* values were determined by Fisher exact testing.

***(F)*** Box plots of genomic dedifferentiation signature[8] (top) and IM4 signature[9] (de-clear cell differentiated (DCCD) ccRCC; bottom) scores between gene-signature-predicted survival ecotypes in the TCGA KIRC cohort. *P* values were determined using Kruskal-Wallis tests.

***(G)*** Heatmap of expression Z-scores of gene signatures of the poor and favorable survival ecotypes derived from H&E prediction and split by NTP-based gene-signature-predicted survival ecotype prediction results in the CPTAC ccRCC cohort[10] (n=212 patients). Patients were assigned to the *Poor* and *Favorable* survival ecotypes based on an FDR threshold (FDR < 0.05) and otherwise called *Medium outcome*.

***(H)*** Kaplan-Meier curves of OS for gene-signature-predicted survival ecotypes in the CPTAC ccRCC cohort (n=195 patients). *P* values were estimated using a log-rank test.

***(I)*** Heatmap of mean expression Z-scores for selected genes across gene-signature-predicted survival ecotypes in the CPTAC ccRCC cohort. Number of patients per group are shown and genes are annotated by their protein-level ecotype association in the discovery cohort.

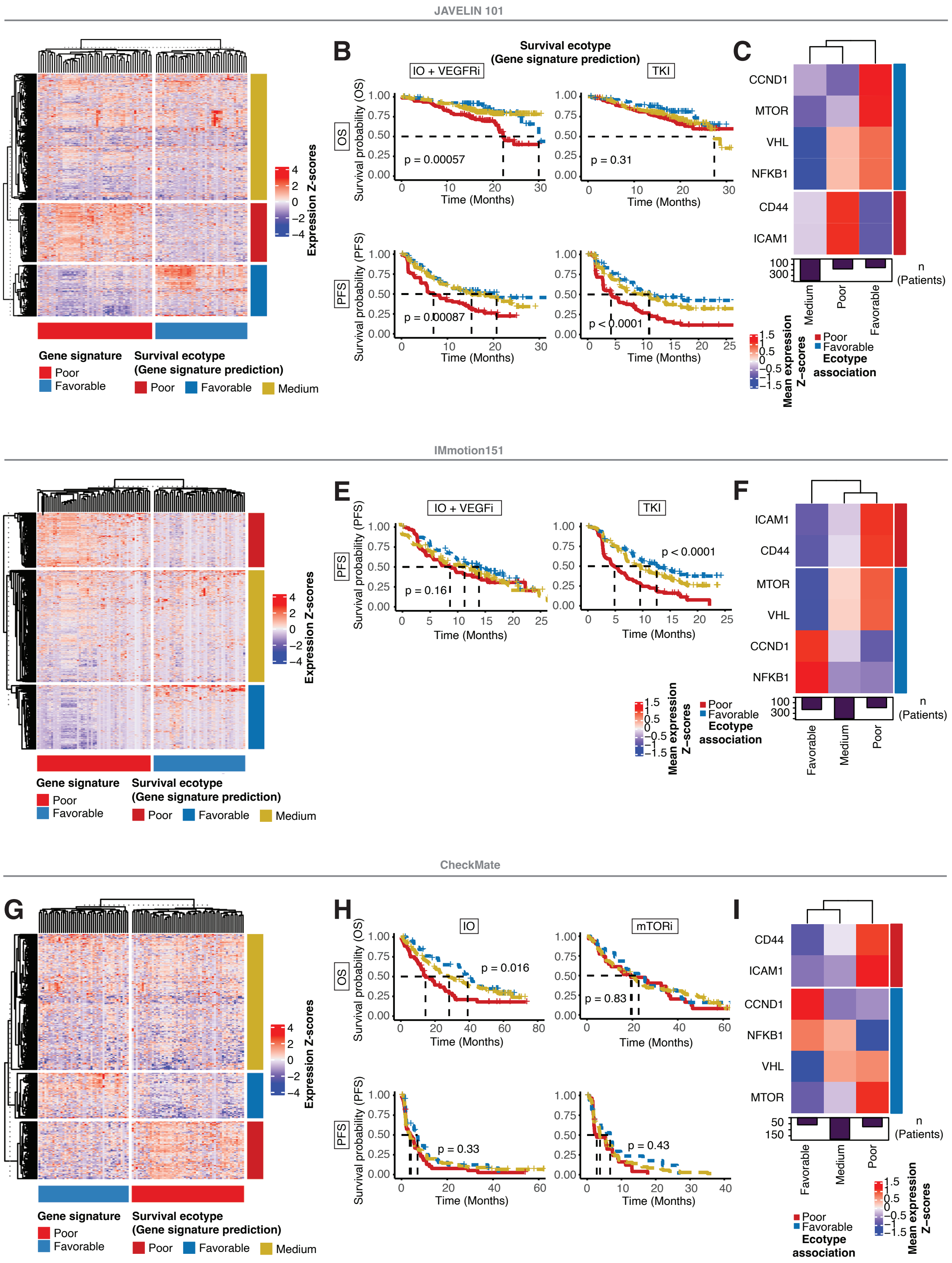
JAVELIN 101
B
Survival ecotype
(Gene signature prediction)
IO + VEGFRi
TKI
OS
Survival probability (OS)
p = 0.00057
p = 0.31
Time (Months)
PFS
Survival probability (PFS)
p = 0.00087
p < 0.0001
C
CCND1
MTOR
VHL
NFKB1
CD44
ICAM1
n
(Patients)
Medium
Poor
Favorable
Mean expression
Z−scores
Ecotype
association
Expression Z-scores
Gene signature
Poor
Favorable
Survival ecotype
(Gene signature prediction)
Medium
IMmotion151
E
F
IO + VEGFi
TKI
p = 0.16
p < 0.0001
CheckMate
G
H
I
IO
mTORi
p = 0.016
p = 0.83
p = 0.33
p = 0.43

**Figure S10: Survival ecotype prediction in clinical trial datasets.** *Related to* ***Figure 6****.*

***(A, D, G)*** Heatmap of expression Z-scores of gene signatures of the poor and favorable survival ecotypes derived from H&E prediction and split by NTP-based gene-signature-predicted survival ecotype prediction results in ***(A)*** the JAVELIN 101 cohort[11] (n=733 patients), ***(D)*** the IMmotion 151 cohort[12] (n=823 patients) and ***(G)*** the CheckMate cohort[13] (n=311 patients). Patients were assigned to the *Poor* and *Favorable* survival ecotypes based on an FDR threshold (JAVELIN 101, IMmotion 151, FDR < 0.05; CheckMate, FDR < 0.1) and otherwise called *Medium*.

***(B, E, H)*** Kaplan-Meier curves of OS and progression-free survival (PFS) for gene-signature-predicted survival ecotypes across treatment arms of ***(B)*** JAVELIN 101, ***(E)*** IMmotion 151 and ***(H)*** CheckMate. *P* values were estimated using a log-rank test.

***(C, F, I)*** Heatmap of mean expression Z-scores for selected genes across gene-signature-predicted survival ecotypes in ***(C)*** the JAVELIN 101 cohort, ***(F)*** the IMmotion 151 cohort and ***(I)*** the CheckMate cohort. Number of patients per group are shown and genes are annotated by their protein-level ecotype association in the discovery cohort.
